# STELLAR: A flexible ensemble learning framework integrating rare variants to enhance polygenic risk prediction

**DOI:** 10.64898/2026.06.07.26355109

**Authors:** Tony Chen, Xihao Li, Haoyu Zhang, Rahul Mazumder, Xihong Lin

**Affiliations:** Department of Biostatistics, Harvard TH Chan School of Public Health, Boston, MA, USA; Department of Biostatistics, University of North Carolina at Chapel Hill, Chapel Hill, NC, USA; Department of Genetics, University of North Carolina at Chapel Hill, Chapel Hill, NC, USA; Division of Cancer Epidemiology and Genetics, National Cancer Institute, Bethesda, MD, USA; Operations Research and Statistics Group, Sloan School of Management, Massachusetts Institute of Technology, Cambridge, MA, USA; Department of Statistics, Harvard University, Cambridge, MA, USA

## Abstract

Whole-exome and whole-genome sequencing technology has enabled the discovery of rare genetic variants associated with human health and diseases. However, existing statistical methods used for rare variant association testing are not well-suited for building genetic risk prediction models that jointly incorporate rare and common variants. We propose STELLAR, a flexible ensemble learning-based approach to compute rare variant polygenic risk scores (PRS) using association summary statistics to enhance conventional common variant PRS. Our method combines burden-based and penalty-based rare variant analysis and leverages functional annotation information to prioritize potentially causal variants within the prediction models. In simulation studies, PRS using STELLAR consistently showed the highest prediction accuracy compared to models using common variants alone or rare variant burdens. Applied to UK Biobank whole-exome sequencing data (n=310,831) across eight continuous and five binary traits, STELLAR significantly improved prediction accuracy, refined stratification of individuals at the highest genetic risk beyond common variants, and prioritized biologically relevant genes. STELLAR provides a scalable strategy to incorporate rare variants into PRS in addition to common variants, advancing precision risk prediction and enabling more comprehensive assessment of genetic contributions to complex diseases.

## Introduction

Polygenic risk scores (PRS) have emerged as powerful tools to translate genetic information into personalized clinical interventions. However, statistical methods to compute PRS have primarily focused on common genetic variants (minor allele frequencies [MAF] more than a specific cutoff like 1% in a population)^1^. Common variants are widely used because they offer sufficient statistical power for useful risk prediction, but they only comprise less than 3% of variation in the human genome^2^. In contrast, rare variants, though only found in a few individuals, can be highly deleterious and lead to extreme health conditions^3–6^.

The emergence of whole-exome sequencing (WES) and whole-genome sequencing (WGS) presents an unprecedented opportunity to identify rare variants contributing to human health and disease^7–10^. Recently-developed statistical models have enabled scalable analysis of such data and led to the discovery of new genetic associations that can inform clinical decisions and drug discoveries^11–13^. The burden test, which collapses rare variants into sets such as genes or functional categories (e.g. loss-of-function, missense), is a popular strategy due to its simplicity^14–16^. However, it makes a strong assumption that all variants within that set have similar effect size and effect direction^17^. While this is often the case for key variant groups such as loss-of-function and disruptive-missense, this hinders the possibility to incorporate genes that exhibit more variable variant-level effect patterns.

To address this, methods such as SKAT^18^ (sequential kernel-based association testing) evaluate variant effects using a variance component test, demonstrating better detection power when effect directions vary within a functional category. However, SKAT can show reduced power when burden assumptions hold. To maximize detection across the genome, the STAAR framework^11^ (variant-set test for association using annotation information) proposes ensemble testing to combine test results from burden, SKAT, and ACAT-V^19^ to powerfully identify gene associations regardless of the genetic effect pattern. Furthermore, STAAR can integrate functional information through multiple variant annotations^20^, which provides more flexibility to prioritize biologically functional variants within a functional category. While this has shown immense promise from an association testing perspective, a limitation is that SKAT does not directly provide effect size estimates that could be used for external risk prediction like PRS.

While methods typically used for common variants can be applied to rare variants^21^, analysis of single rare variants has limited power and the number of rare variants is much larger than the number of common variants, making it difficult to apply common-variant-based PRS methods to rare variants. Thus, existing efforts to integrate rare variants have largely relied on burden-based scores, using collapsed effect sizes for each variant set^5,22,23^. Disease-specific applications have shown that rare variant burden scores can improve risk prediction for coronary artery disease^24^, type 2 diabetes^25^, and autism spectrum disorder^26^, and can capture highly penetrant effects overlooked by common-variant PRS. More recently, multi-ancestry PRS framework RICE^22^ integrated burden-based rare variant PRS derived from STAARpipeline^12^ with common variant PRS, enabling systematic evaluation across multiple traits and ancestries. Together, these studies highlight the potential of rare variant burden scores for prediction, but also their limitations: reliance on restrictive effect assumptions at the set level and limited flexibility to model variant-level heterogeneity.

Thus, there remains a critical need for new statistical methods that can (1) flexibly model rare variant effects beyond the burden model and (2) leverage gene structure and functional annotations to maximize prediction power from rare variants. To address this, we propose STELLAR (summary-<u>ST</u>atistics based <u>E</u>nsemb<u>L</u>e <u>L</u>earning with functional <u>A</u>nnotations for <u>R</u>are variant PRS), a fast ensemble-based method to incorporate rare variants in PRS. STELLAR integrates burden- and penalty-based rare variant analyses with functional annotation information for flexible, biologically informed risk prediction. Through extensive simulations and applications to eight continuous and five binary traits using WES data in the UK Biobank^10^ (n=310,831), STELLAR achieved improved prediction accuracy, enhanced risk stratification, and biologically interpretable gene prioritization compared with common variant PRS (cvPRS) and burden-based rare variant PRS (rvPRS). These features highlight our method’s potential clinical utility in identifying individuals at the highest risk who will most likely benefit from targeted interventions.

## Results

### Method overview

STELLAR is a scalable ensemble learning framework for building risk prediction models from rare variants (**Figure 1**). Using summary-level rare variant association statistics and linkage disequilibrium (LD) references, STELLAR fits three complementary base learners: (1) burden, (2) single-variant L0L2 penalized regression ^27,28^, and (3) group L0L2 penalized regression^29^ applied to variant sets for group-level selection and within-group shrinkage. The final ensemble step combines these three learners via cross-validation to accommodate both same-direction and mixed-direction effects.

**Figure 1.**
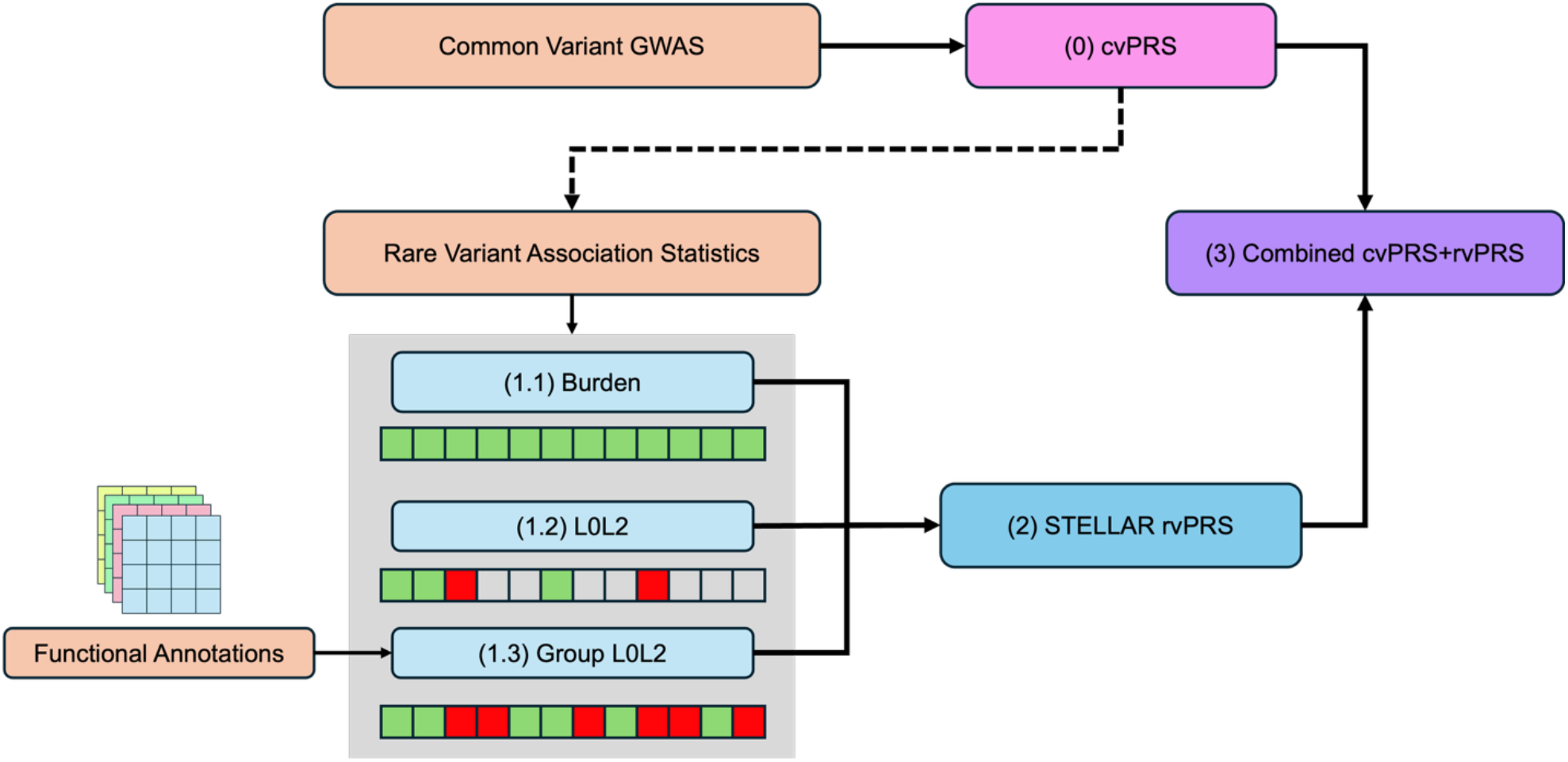
Overview of the STELLAR framework. The core of STELLAR comprises three models: (1.1) burden, which assumes effect sizes are consistent within a mask; (1.2) single-variant L0L2 penalized regression; and (1.3) group L0L2 penalized regression, which can be supplemented by functional annotation information. (2) We then use ensemble learning to combine candidate models into a rare variant PRS (STELLAR rvPRS), which can be used in conjunction with (0) a common variant PRS (cvPRS) using methods such as ALL-Sum on common variant GWAS summary statistics to provide a (3) final combined PRS with both common and rare variants.

We defined variant sets or “masks” using gene positions and variant-level functional categories: loss-of-function (LOF), disruptive missense (DMIS), missense (MIS), synonymous (SYN), promoter (PRO), enhancer (ENH), downstream (DOWN), upstream (UP), and untranslated region (UTR)^12^. This grouping can improve detection power when rare variants and their signals are too sparse to analyze individually. Like in STAAR, STELLAR can also incorporate functional annotations to prioritize predicted functional variants by down-weighting their penalties, while imposing stronger penalties on variants with lower predicted functional relevance. The resultant rvPRS can be combined with cvPRS to yield a joint score, allowing rare variation to complement common-variant prediction.

From the ensembled rvPRS, we also derived importance metrics based on variant-level effect estimates in each gene mask. These metrics provide structured interpretation of the PRS by identifying gene masks that contribute most to disease risk, facilitating deeper biological and translational investigation of risk-associated genes.

### Simulation studies

To benchmark STELLAR, we simulated phenotypes using observed UKBB genotype and WES data on chromosome 19 (**Methods**). We analyzed 127,820 participants, splitting 97,820 for training to compute common and rare variant association summary statistics, 15,000 tuning samples to conduct PRS ensembling, and 15,000 validation samples to evaluate prediction accuracy. For common variants, we included 32,947 variants based on the HapMap3^30^ and MEGA^31^ arrays. The WES data for chromosome 19 included 563,126 total rare variants; after collapsing ultrarare variants (MAC < 10) within each mask^13,19^, 110,447 effective rare variants remained. We also generated five synthetic functional annotation weights from a standard multivariate normal distribution.

We tested a wide range of simulation settings, generating 50 sets of continuous phenotypes under each setting. We randomly selected 10% of the common variants to be causal with effect sizes drawn from a normal distribution corresponding to a heritability of 5%. Then, for the rare variants, we varied the number of causal genes (three or five), genetic effect sizes (small effects *β* = 0.15|*MAF*| or large effects *β* = 0.25|*MAF*|), effect size directions within each mask (100%, 80%, or 50% in the same direction), and proportion of causal rare variants within each mask. The causal status of rare variants was determined by their gene mask (LOF, MIS, or SYN) and their functional annotation weights (see **Methods**). We also evaluated additional settings where ultrarare variants were about twice as likely to be causal. We compared three sets of PRS models: (1) cvPRS using the ALL-Sum method^27^, (2) a combination of cvPRS and Burden-based rvPRS (cvPRS+Burden), and (3) a combination of cvPRS and STELLAR-based rvPRS (cvPRS+STELLAR). Our primary metric was the prediction R^2^ in the 15,000 validation samples, averaging across the 50 simulations under each simulation setting.

Across all simulation settings, cvPRS+STELLAR achieved the highest R^2^, with consistently significant improvements over cvPRS alone and similar or significantly higher R^2^ than cvPRS+Burden (**Figure 2, Supplementary Table 1**). In particular, under the settings where the rare variant effects were 50% positive and 50% negative, cvPRS+Burden showed limited improvement compared to cvPRS (2% R^2^ increase on average), while STELLAR maintained high accuracy compared to both cvPRS and cvPRS+Burden (14% and 12% R^2^ increase on average, respectively) (**Supplementary Figure 1A-B**). With three causal genes and strong rare variant effects (*β*_*j*_ = 0.25|*MAF*_*j*_|), STELLAR increased the R^2^ by 8% (p<0.01) compared to cvPRS and outperformed Burden model by up to 6.5% (p<0.01), even when rare variant effects were all in the same direction. While Burden can be advantageous for consistent effect directions, it still faces challenges from variation in effect sizes and potential sparsity within masks, both of which can be captured through penalized regression. With five causal genes and weaker rare variant effects (*β*_*j*_ = 0.15|*MAF*_*j*_|), cvPRS+STELLAR achieved up to 18.4% higher R^2^ than cvPRS, and 17.4% over cvPRS+Burden.

**Figure 2.**
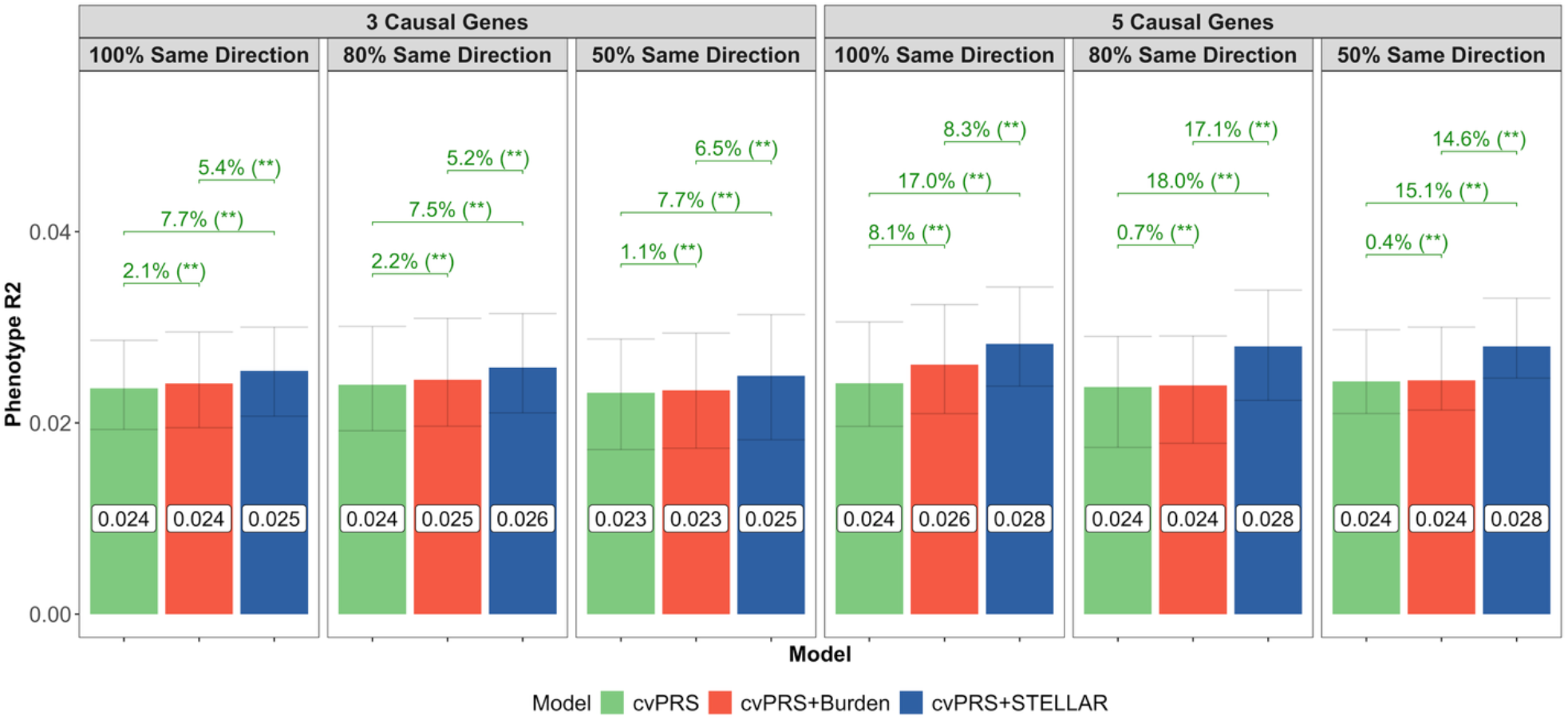
Simulation prediction results across varying causal genes and effect sizes. Using observed genotypes from UK Biobank, we simulated continuous phenotypes with 10% of common variants contributing 5% heritability and varied effect sizes and directions for rare variants. Here, we show two sets of results: (1) 3 causal genes with effects ***β* = 0. 25**| **log**_***1*0**_ ***MAF*** | and (2) 5 causal genes with effects ***β* = 0. 15**| **log**_***1*0**_ ***MAF*** |, with 30% of LOF, 10% of MIS and 3% of SYN rare variants being causal. Summary statistics were computed from 97,820 UK Biobank samples, PRS were tuned using 10,000 samples and validated in a separate 10,000. We compared a baseline PRS with only common variants (CV) using ALL-Sum, a combined PRS of ALL-Sum and Burden scores, and our proposed combination of ALL-Sum and STELLAR. We repeated each simulation setting 50 times, and reported the average R^2^ for each PRS (text boxes) as well as percentage differences between PRSs with Bonferroni-adjusted p-values for these differences using paired t-tests (floating text), where (**) indicates p < 0.01.

Under weak-effect settings with three causal genes, cvPRS+STELLAR significantly improved R^2^ by 1% on average compared to cvPRS. cvPRS+STELLAR also improved R^2^ by 0.5% compared cvPRS+Burden, but the differences were often not significant due to weak rare variant effects (**Supplementary Figure 1A**). With five causal genes and strong effects, cvPRS+STELLAR significantly improved R^2^ by 36% compared to cvPRS and 23% compared to cvPRS+Burden on average (**Supplementary Figure 1B**). In additional simulations where ultrarare variants were more likely to be causal, cvPRS+STELLAR remained the best model across all settings (**Supplementary Figure 1C-D**).

To benchmark the robustness of cvPRS+STELLAR to extraneous annotations, we further compared an implementation using the five informative annotations used to determine the causal status of rare variants with two additional random annotations (**Supplementary Table 2**). Across 48 settings, we observed no systematic change in accuracy, indicating that STELLAR maintains high rvPRS prediction accuracy even with the inclusion of non-informative annotations.

Furthermore, using the importance metrics to evaluate selected masks, we verified that STELLAR primarily prioritized causal masks within the prediction model (**Supplementary Figure 2**). In our simulations, causal MIS masks generally included more variants than LOF masks. As a result, Total Contribution assigned more importance to MIS masks, while the Average Contribution gave more importance to LOF masks. In summary, our simulations show that STELLAR improves overall PRS accuracy across diverse rare variant architectures, while remaining robust to functional annotations and recovering relevant gene masks through its importance metrics.

### STELLAR significantly improves risk prediction accuracy

We applied the three PRS methods to the full UK Biobank with 310,831 unrelated samples of European ancestry for prediction of eight continuous traits (**Figure 3, Supplementary Table 3**): low-density lipoprotein cholesterol (LDL), high-density lipoprotein cholesterol (HDL), log-triglycerides (logTG), total cholesterol (TC), height, body mass index (BMI), estimated glomerular filtrate rate (EGFR), hemoglobin A1C (HbA1c). We divided the UKBB samples into 3 independent datasets: 270,831 training samples to compute rare variant association statistics, 20,000 samples for PRS tuning, and 20,000 for out-of-sample validation. For the cvPRS, we used published consortium-based GWAS summary statistics to compute the PRS and included up to 1,546,685 common variants from the HapMap3^30^ and MEGA^31^ (multi-ethnic genotyping array) SNP panels. We used WES from the UK Biobank for rare variant analysis, which included a total of 8,580,612 rare variants, and was reduced to 1,567,289 effective variants after collapsing ultrarare variants with minor allele count (MAC) less than 10 within each gene mask. For the weighted group L0L2 penalty, we included an unweighted penalty as well as weightings from 11 functional annotations derived from existing scoring software as well as annotation principal components (PCs) from the STAAR paper (**Methods, Supplementary Table 3**). We then evaluated the prediction accuracy using partial R^2^ of cvPRS and rvPRS, adjusting for baseline covariates of age, sex, and the first ten pre-computed ancestry PCs in UKBB. cvPRS+STELLAR significantly increased partial R^2^ by up to 8.6% compared to ALL-Sum cvPRS alone, and up to 6.1% compared to cvPRS+Burden (**Figure 3**). This highlights that STELLAR captures more signal from rare variants than Burden models across a wide range of traits. Taking R^2^ as an approximation of trait heritability, our results show that rare variants can account for as much as 2% of phenotypic variation for height (7.8% of the total 0.257). On the other end, BMI showed much less rare variant signal, about 0.1% of overall phenotypic variation, which is only 1% of the total 0.086.

**Figure 3.**
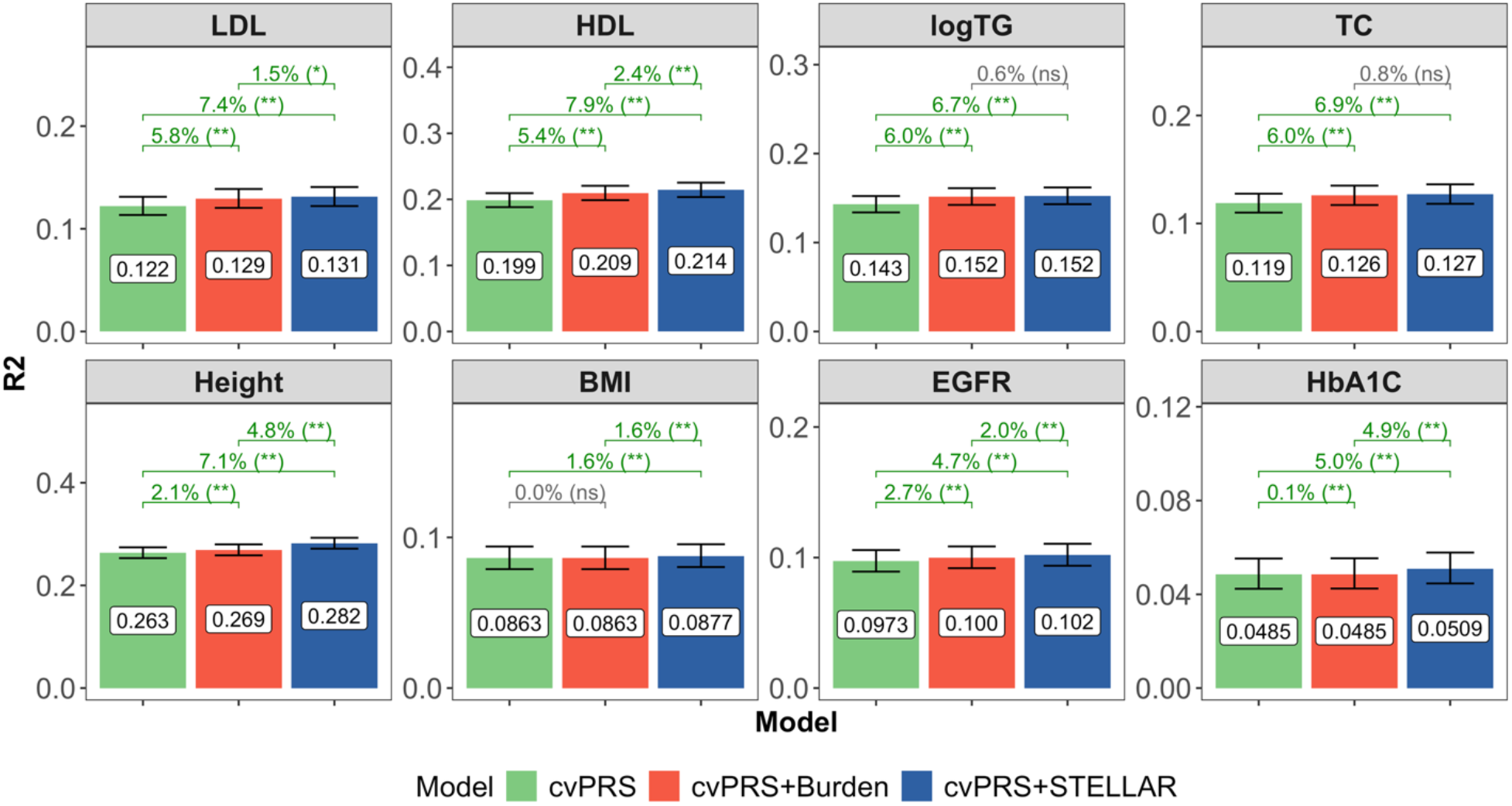
Comparison of R^2^ between PRS models for 8 continuous traits in UK Biobank. We evaluated the three PRS approaches in real continuous traits from the UK Biobank. We used published GWAS summary statistics (n≈130,000 to 930,000) to compute the common variant PRS using ALL-Sum, and computed rare variant association statistics in UK Biobank WES (n≈270,000). We tuned each method in 20,000 UK Biobank samples and validated in an independent 20,000. We show the observed partial R^2^ of each PRS with error bars representing the 95% BCI of partial R^2^. We also evaluated the BCI in the percentage difference in R^2^ between methods, where (**) indicates that the 99% BCI in percentage difference does not include 0, (*) that the 95% BCI does not include 0, and (ns) that the 95% BCI does include 0.

In addition to prediction of continuous traits, we also applied our method to analysis of five binary traits – breast cancer, prostate cancer, coronary artery disease (CAD), type 2 diabetes (T2D), and asthma (**Supplementary Figure 3**). Overall, all three PRS models achieved very similar partial AUCs, adjusting for baseline covariates and excluding sex for breast and prostate cancers. cvPRS+STELLAR showed a significant increase in partial AUC of 0.18% over both cvPRS and cvPRS+Burden for T2D, and an increase of 0.02% over cvPRS and 0.01% over cvPRS+Burden for breast cancer. However, cvPRS+STELLAR did not show any significant improvements for CAD, prostate cancer, or asthma.

We further benchmarked the performance of STELLAR across different combinations of functional annotations, from a single-variant L0L2 model with no functional model up to the full STELLAR model in our main analysis (**Supplementary Figure 4**). Across all traits, we observed that leveraging the group L0L2 penalty with gene and mask structure accounted for the largest improvements in accuracy from a single-variant penalty, with improvements as large as 4% for HbA1C. On top of this, incorporating functional annotation weights within the group penalty provided complementary accuracy gains, with significant improvements in R^2^ of 0.8% for Height and 0.4% for HDL. Of note, incorporation of more functional annotation weights only improved or maintained similar accuracy. This reinforces that STELLAR is robust to potentially uninformative annotations, making it flexible to incorporate information from diverse biological resources. Therefore, annotations from new technologies and methods could also be used in future studies to develop the best prediction model for a trait. Owing to the scalability of penalized regression frameworks, we found that all-chromosome analysis could be completed within one hour for any annotation and about 10GB of memory, so the overall analysis scales approximately linearly with the number of input annotations.

### Adjusting for common variants enhances risk prediction power from rare variants

In our primary analysis framework (**Figure 1**), we used cvPRS as a covariate in rare variant association testing. This approach has been shown to enhance power to detect rare variant signals^32^ and reducing potential confounding from nearby common variants^12,33^. However, cvPRSs are not always readily available for new rare variant association studies. To assess the impact of this adjustment, we compared STELLAR’s prediction accuracy with and without adjusting for cvPRS during rare variant association analysis (**Supplementary Figure 6**). Across all eight continuous traits, both implementations of STELLAR significantly outperformed cvPRS, demonstrating the robust predictive value of rare variants. However, adjusting for common variants further increased STELLAR’s prediction, with R^2^ gains of 0.7% for HDL, 0.8% for LDL, and 1.3% for Height.

To better understand these gains, we examined rare variant association results on chromosome 19 for LDL (**Supplementary Figure 7**). Of the 111,249 single-variant and ultra-rare-burden associations tested, 111,228 (99.98%) were not significant (p > 5×10^−8^) regardless of PRS adjustment. A small number of associations changed status: one variant became significant only after adjustment, 11 lost significance, three became less significant, and six became more significant. These results indicate that adjusting for PRS primarily removes overlapping signals between rare and common variants, while enhancing detection of independent rare variant effects.

Five of the six variants with more significant associations were mapped to *LDLR*, which has well-established roles in LDL. This is likely because including cvPRS reduces residual variation in the outcome, therefore enabling even stronger associations from these variants whose signals are not captured by cvPRS. Altogether, these findings show that whenever possible, rare variant association testing should adjust for common variant PRS to both remove redundant effects and strengthen independent rare variant signals.

### STELLAR refines individual risk stratification beyond common variants alone

While metrics such as R^2^ and AUC are the most commonly used to evaluate PRS performance, they only characterize population-level correspondence between predicted and true outcomes. As noticeable in our results, these metrics show fairly small absolute differences between methods, as most individuals in the data do not carry risk-associated rare variants, and thus the contribution of rare variants to R^2^ and AUC may get diluted. To assess STELLAR’s ability to refine individual-level risk, we examined stratified risk profiles across the common and rare variant PRS distributions.

For continuous traits, we quantified relative risk as the standardized phenotypic difference between extreme PRS quantiles and a median reference group, adjusting for baseline covariates (age, sex, and the first ten ancestry PCs) (**Figure 4, Supplementary Figure 8**). Using a global test for stratification (**Methods**), STELLAR rvPRS showed significantly improved risk stratification beyond cvPRS in all traits except for BMI at the bottom 10% of rvPRS, highlighting the value of rvPRS in refining individual-level risk. Among individuals in the top 10% of both cvPRS and rvPRS, the average relative risk was 0.708 (s.d.=0.207), 25% higher than the top 10% of cvPRS (mean=0.568, s.d.=0.131). Similarly, the bottom 10% of both PRSs yielded a relative risk of -0.740 (s.d.=0.132), which was 27% lower than cvPRS alone (−0.582, s.d.=0.116).

**Figure 4.**
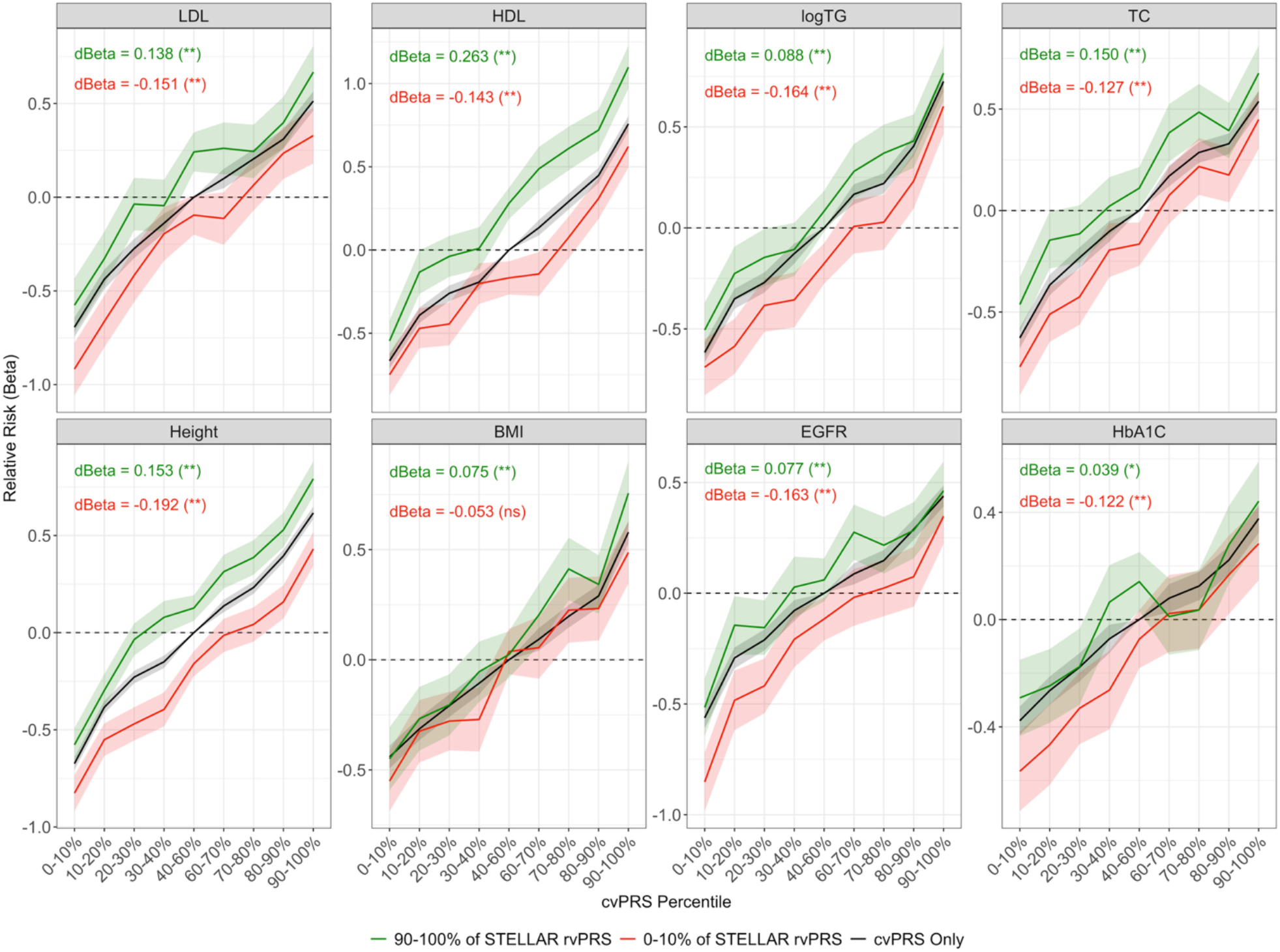
Individual-level risk stratification for continuous traits in UK Biobank. We stratified individuals in the UK Biobank validation dataset (n≈20,000) according to their percentiles of the common variant (ALL-Sum) and rare variant (STELLAR) PRS distributions. For the common variants, we calculated the relative risk (Beta, y-axis) for each continuous trait using linear regression with each percentile as a categorical variable (40-60% as the reference group) and the outcome standardized to have mean 0 and variance 1. We also calculated relative risks according to both common and rare variants, using 40-60% in both distributions as the reference group. The black line denotes the relative risk using only cvPRS, and the green and red lines show the relative risk for each cvPRS further stratified by the top and bottom 10% of the STELLAR rvPRS, respectively. Ribbons correspond to the 95% confidence intervals for each Beta estimate. Text in the top left corner shows the average difference in Beta (“dBeta”) between the top 10% of STELLAR rvPRS and cvPRS (green) and the bottom 10% of STELLAR rvPRS and cvPRS (red), with (**) for p < 0.01, (*) for p < 0.05, and (ns) for non-significant differences from one-sided tests (see **Methods**).

For the five binary traits, we evaluated the odds ratio (OR) of disease relative to the median PRS group, adjusting for baseline covariates (excluding sex for breast and prostate cancers) (**Supplementary Figure 9**). While the global test for stratification showed no statistically significant differences between cvPRS only and cvPRS+STELLAR stratified risk, higher rvPRS was still associated with increased risk for disease. Individuals in the top 10% of both cvPRS and rvPRS showed an average OR of 3.06 (s.d.=1.20), 15% higher than the top 10% of cvPRS (mean OR=2.66, s.d.=0.780).

Overall, these results demonstrate that STELLAR not only enhances population-level prediction accuracy over cvPRS but also enables more fine-grained individual-level risk stratification, which can help identify the individuals at the highest genetic risk who may benefit most from personalized interventions.

### Rare variants improve prediction accuracy across biobanks and genetic ancestries

We further evaluated prediction accuracy of the cvPRS and cvPRS+STELLAR models, all trained in UKBB European ancestry, across six ancestry groups (**Figure 5, Supplementary Table 8**). In addition to the main validation cohort, individuals of European (EUR) ancestry with prediction ancestry probability above 90% (**Methods**), we also assessed prediction in individuals for whom European ancestry was the most likely label but had predicted ancestry probability less than 90%, along with individuals predicted as Admixed American (AMR), South Asian (SAS), East Asian (EAS), and African (AFR) ancestry. Across all non-European ancestries, while we still observed lower R^2^ relative to European ancestry, incorporating STELLAR rvPRS reduced the overall decline in accuracy and showed significant improvements over cvPRS alone. Averaged over the 8 continuous traits, the relative R^2^ for EUR with probability less than 90% was 0.811 when only using common variants, but increased up to 0.876 (8.4%) with STELLAR rvPRS (p < 0.01). We also saw increases in R^2^ by 7.9% in AMR (p = 0.04), 5.4% in SAS (p = 0.02), 6.1% in EAS (p = 0.01), and 14.3% in AFR (p = 0.09). While the improvements in African ancestry were not statistically significant, this may also be driven by trait-specific variability in accuracy. For example, accuracy was nearly 0 in HbA1C but actually higher than in other non-European ancestries for LDL and TC (**Supplementary Figure 10**).

**Figure 5.**
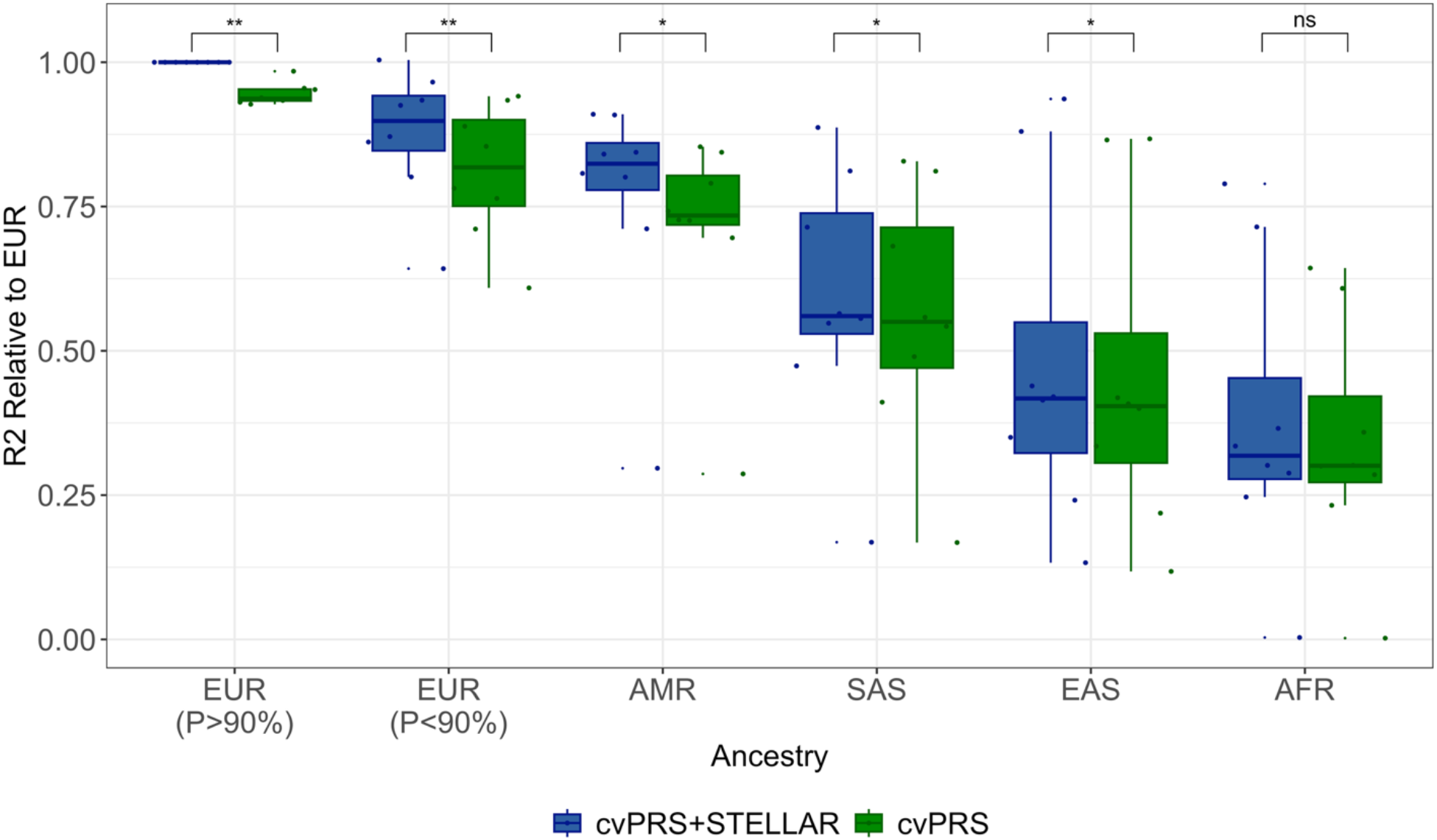
Prediction accuracy across ancestry groups in UK Biobank. We evaluated PRS prediction accuracy for UKBB samples of non-European predicted ancestry (n≈24,000) and individuals of European predicted ancestry with PC-predicted ancestry less than 90% (n≈5,000). We compared the relative accuracy of each PRS compared to cvPRS+STELLAR in European ancestry with PC-predicted ancestry greater than 90%, which is most similar to the UKBB training and tuning samples. Points and boxplots indicate relative R^2^ for each of the 8 continuous traits, colored by either cvPRS (green) or cvPRS+STELLAR (blue). We used paired t-tests to compare relative accuracy within each ancestry: (**) p < 0.01; (*) p < 0.05; (ns) p > 0.05.

We conducted further multi-ancestry validation in 228,954 unrelated individuals from the All of Us (AoU) cohort^34^, with 100,775 (44%) from non-European ancestry. We evaluated the same UKBB-based cvPRS and cvPRS+STELLAR PRSs in six ancestry groups: EUR, MID (Middle East), AMR, SAS, EAS, and AFR. Across all ancestry groups, we saw the same improvements in prediction accuracy (**Supplementary Figure 11**). Compared to cvPRS only, cvPRS+STELLAR yielded 5.8% higher R^2^ in EUR (p < 0.01), 21.9% in MID (p = 0.02), 3.4% in AMR (p < 0.01), 3.3% in SAS (p < 0.01), 2.7% in EAS (p = 0.03), and 80.3% in AFR (p = 0.08).

In summary, rare variant effects estimated by STELLAR are significantly predictive even when trained in European ancestry and applied to non-European ancestries, which was robustly verified in both UKBB and AoU.

### STELLAR prioritizes biologically relevant genes through importance metrics

In addition to improving risk prediction and stratification, STELLAR also identifies biologically relevant genes for each of the analyzed traits via importance metrics (**Supplementary Figure 12, Supplementary Figure 13**). For instance, in analysis of lipid traits, STELLAR assigned high importance to well-established genes such as *LDLR, PCSK9, LCAT*, and *CETP*^35,36^. In addition to correctly prioritizing well-known genes, STELLAR also importantly included masks such as *LDLR* loss-of-function variants that were not even nominally significant in burden tests. Further dissection of our results showed that the variant-level effect sizes within this mask included both large positive and large negative effects, which would cancel out in a simple burden model, demonstrating the valuable flexibility offered from the penalized regression models (**Supplementary Figure 14, Supplementary Figure 15**). STELLAR also prioritized loss-of-function and disruptive missense variants on genes such as *ACAN, ADAMTS10*, and *DTL* for height^37^; *CTRC1* and *LTA* for EGFR, both related to chronic kidney disorder^38,39^; and *APPBP2* for HbA1c, which could be involved in pathways for T2D^40^. On the other hand, the masks prioritized for BMI were all identified as noncoding (e.g. untranslated regions or promoter regions) and showed no clear evidence of association with BMI, but this coincides with the minimal observed contribution of rare variants to R^2^ (**Supplementary Figure 6**).

For traits like Height and HDL, many of the high-importance masks appear to be driven by ultrarare variants, as the estimated effect sizes were concentrated at either single points or very narrow boxplots (**Supplementary Figure 14, Supplementary Figure 15**). This demonstrates that by separating variants within a mask, STELLAR can more appropriately pinpoint which sections of the genome contribute the most to disease. On the other hand, while burden does select many of the same masks, it can only nominate an entire mask, which cannot provide information for individual variants or delineate between rare versus ultrarare variants.

In the analysis of disease traits, even though prediction and stratification improvement were not as noticeable as the continuous traits, STELLAR still prioritized several relevant genes that were not selected by the Burden ensemble model. For example, STELLAR assigned high importance to genes such as *GCK* for T2D^41^, *LDLR* for CAD^42^, *BRCA2* and *PALB2* for breast cancer^43,44^, and *SIRT1* for prostate cancer^45^. There were also several other genes without well-established connections to disease. For example, the highest importance for breast cancer was given to the loss-of-function variants on the *RGL3* gene. This gene is related to the RAS pathway commonly associated with cancer^46^, but has only recently been studied in relation to breast cancer^47,48^. This demonstrates that STELLAR can provide additional support for investigation of previously under-studied genes and deeper insights into disease mechanisms.

Together, these results demonstrate that STELLAR-derived importance metrics can complement association testing by prioritizing biologically relevant, risk-associated gene masks. This provides an interpretable link between overall risk prediction and disease mechanisms that may inform future functional and translational studies.

## Discussion

In this work, we introduced STELLAR as a statistical method to incorporate functional annotations and rare variants within PRS models. STELLAR combines burden models and penalized regression through an ensemble learning approach that flexibly models rare variant effects. Through comprehensive simulation studies, we showed that a combined cvPRS+STELLAR model significantly outperforms both cvPRS and cvPRS+Burden models, especially when rare variant effect directions varied within masks. In our analysis of 13 traits and diseases in UKBB, cvPRS+STELLAR consistently yielded the highest R^2^ across all methods. We observed that leveraging gene-mask structure in the group L0L2 penalty was the primary driver of STELLAR’s improved prediction accuracy, and additionally incorporating functional annotations as weights can contribute meaningful gains for traits such as Height and HDL. Importantly, STELLAR is robust across different combinations of annotation weights and can thus be used to integrate trait-specific, multi-omic, and deep learning-derived annotations to further improve prediction accuracy^49–53^.

Additionally, STELLAR enabled sharper individual-level risk stratification, particularly at the extremes of the risk distribution, suggesting potential clinical utility of rare variants in identifying individuals at elevated genetic risk^5^. While R^2^ and AUC are still valuable metrics to evaluate PRS, quantile-based metrics can offer deeper insights into the clinical utility of rvPRS, especially when considering the small population-level contribution to risk from rare variants and the ultimate goal of identifying individuals who will benefit most from personalized interventions.

Beyond predictive performance, STELLAR also provides importance metrics to prioritize gene masks and elucidate trait-relevant biology. For example, STELLAR correctly prioritized known genes such as *PCSK9* and *BRCA2*, while also identifying novel candidates like *RGL3* for breast cancer. These results demonstrate that STELLAR can complement standard association testing by nominating both established and previously understudied loci for follow-up. In traits like Height and HDL, STELLAR prioritized masks driven by ultra-rare variants, where standard burden models may fail due to effect size heterogeneity. Importantly, STELLAR was also able to prioritize LOF variants within *LDLR*, which were not selected by the burden-based approach, that showed both positive and negative effects.

Furthermore, STELLAR demonstrated consistent prediction accuracy gains across diverse genetic ancestries in UKBB and AoU, showing significantly higher accuracy in AMR, EAS, SAS, and MID, with large near-significant improvements in AFR. However, because the PRSs were trained in EUR samples, accuracy in non-EUR ancestries remained lower than in EUR. This underscores the need to expand diversity in WGS data, as well as appropriate methodology to model global genetic heterogeneity to ensure optimal risk prediction and equitable clinical use of PRS across all populations^22,54–56^.

Although prediction of binary traits was modest overall, STELLAR nevertheless improved AUC for T2D and breast cancer, refined stratification for individuals at extremely high genetic risk, and prioritized known disease genes like *BRCA2* (breast cancer), *GCK* (T2D), and *SIRT1* (prostate cancer). We expect that future expansion of WES and WGS data in disease-focused consortia^57–59^ will advance both prediction accuracy and biological insights. Additional method refinements to use summary statistics from approaches like SAIGE or REGENIE to correct for case-control imbalance^13,60,61^, MetaSTAAR and MultiSTAAR that combine multiple studies or multiple traits^62–64^, and other regression models could also enhance overall prediction modeling^28^.

We note several limitations and areas for future development. First, STELLAR requires individual-level data to adjust for cvPRS in rare variant association testing and to conduct ensemble learning, which limits its applicability in contexts when only summary statistics are accessible. Although methods for tuning PRS using only summary statistics are available^65–69^, they are primarily designed for common variants and may not translate directly to rare variant analysis. Extending STELLAR to operate entirely on summary statistics, particularly for rare variants, will be a key future direction. Second, STELLAR models common and rare variants separately, using cvPRS to adjust rare variant association testing. While this modular framework simplifies computation, future methods to jointly model common and rare variants could improve both prediction accuracy and biological interpretation. Finally, our study focused on WES data, which captures a subset of coding and functional variation. While collapsing ultra-rare variants helped reduce model dimension, our framework will require scalable extensions to accommodate WGS data, especially to capture noncoding variants that may explain remaining rare variant heritability^71^.

In conclusion, STELLAR offers a scalable framework for integrating rare variants and functional annotations into polygenic risk prediction. By flexibly modeling rare variant effects, STELLAR improves prediction accuracy, refines risk stratification, and yields interpretable insights into disease biology. These features lay important groundwork for expanding PRS beyond common variation toward more comprehensive genetic risk profiling and ultimately more accurate precision medicine.

## Data Availability

Software and tutorials are available at https://github.com/chen-tony/STELLAR. UK Biobank genotype and phenotype data were obtained through application no. 52008. Data from 1000G were obtained from https://www.cog-genomics.org/plink/2.0/resources (genotypes) and https://www.internationalgenome.org/data-portal/sample (population labels). Published GWAS summary statistics for common variants are listed in Supplementary Table 3D.

## Methods

### STELLAR model components

STELLAR takes an ensemble of three different prediction models for rare variants: burden-based model, single-variant L0L2 penalized regression, and group L0L2 penalized regression. *Common variant PRS* were computed using ALL-Sum^27^, which leverages a L0L2-penalized regression model from common variant association statistics *r*^*CV*^ and a block-diagonal LD matrix *R*^*CV*^, defined using pre-defined LD hotspots. For variant *j*, 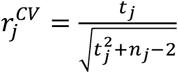 corresponds to 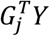 on a standardized scale, where *t*_*j*_ is the test statistic from marginal regressions of the outcome *Y* on the genotype *G*_*j*_, and *n*_*j*_ is the observed sample size. Likewise, the LD is computed as the correlated between variants within each gene, corresponding to 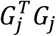 on the standardized scale.

The penalized regression model is given as

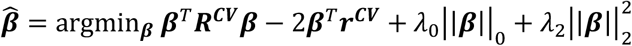

where ∣|*β*|∣_0_ = ∑_*j*_ *l*(*β*_*j*_ ≠ 0) is the L0 norm penalty with tuning parameter *λ*_0_ to control model sparsity, and 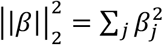 is the L2 norm penalty with tuning parameter *λ*_2_ for effect size shrinkage. This penalized regression can be efficiently solved using cyclical coordinate descent, which iterates through each variant *j* one at a time until convergence. For a particular iteration *t* with current effect estimates ***β***^(*t*−1)^:

<u>Step 1</u>: Compute working correlations: 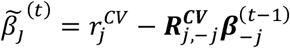

<u>Step 2:</u> Apply penalty: 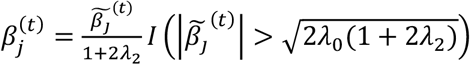

These estimates are then ensembled using 3-fold cross-validation lasso within an independent tuning dataset to compute common variant PRS *PRS*_*CV*_.

*Burden* models are based on computing “burdens” for each gene and functional category (or “masks”). To define these masks, we first used known gene locations from the STAARpipeline package version 0.9.7^76^ and extract genotypes from WES data in aGDS format^20,77^. Variants located between genes were labeled as “Intergenic”, with one “Intergenic” region between each pair of genes. Then, functional categories were defined using annotations in the aGDS dataset following the STAARpipeline9,14: loss-of-function (LOF), disruptive missense (DMIS), missense (MIS), synonymous (SYN), promoter (PRO), enhancer (ENH), downstream, (DOWN), upstream (UP), and untranslated region (UTR). For each mask, we took the row sum of the genotype matrix, where any missing genotype values were imputed with the overall mean. This corresponds to the total number of minor alleles that an individual carries across all variants within that mask. Specifically, if *G* is the full *n* × *d* genotype matrix (with *n* individuals and *d* variant) and *V*_9_ is the index set of variants in mask *m*, the burden for an individual *i* and mask *m* is computed as 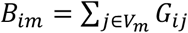. We then performed marginal regression analyses of each burden with the residuals of the outcome:

<u>Step 1</u>: Regress *Y* ∼ *PRS*_*CV*_ + covariates

<u>Step 2</u>: Compute residuals *res* = *Y* − *Ŷ*, where *Ŷ* are the fitted values from Step 1

<u>Step 3</u>: Marginal regressions: *res* ∼ *β*_*m*_*B*_*m*_ where *β*_*m*_ is the marginal effect of *B*_*m*_

In Step 2, the fitted values are computed using linear regression and logistic regression for disease traits. Then, in Step 3, *res* are always continuous values for which we use linear regression to conduct marginal associations. From this, we can derive marginal association p-values *p*_*m*_ for each mask, and compute candidate burden prediction models by p-value thresholding. That is, for a p-value threshold 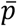, the corresponding burden prediction *S*_*i*_ for an individual *i* is calculated as

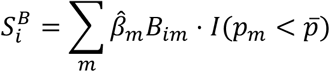

where 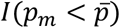 is an indicator that takes value 1 if the p-value *p*_*m*_ falls below 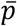. In our analyses, we considered values 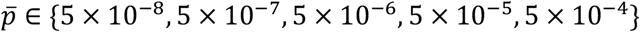.

*Single-variant L0L2* utilizes single-variant association statistics rather than aggregated burden association statistics. Similar to ALL-Sum, the association statistics are computed by marginal regressions of the residuals *res* (i.e. the outcome adjusted for the common variant PRS and covariates) on each variant *j*. Then, the values 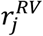 correspond to 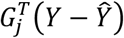 on a standardized scale, similar to the score statistics typically used in rare variant association testing. Using a gene-based block-LD matrix ***R***^***RV***^ among the rare variants, the penalized regression is given as

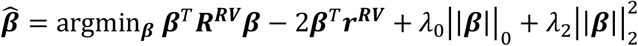

*Grouped L0L2* is conducted similarly using single-variant association statistics and LD, but applies the L0L2 penalty on masks rather than single variants:

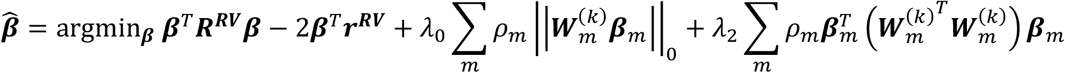

where 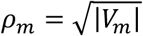, or the square-root of the number of variants in mask *m*, such that the penalty can be applied more evenly across both large and small masks. ***W***^(*k*)^ is a diagonal matrix of weights that can be derived based on functional annotation *k*, and 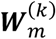 is the submatrix corresponding to mask *m*. For an unweighted implementation, we take 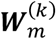 to be the identity matrix ***I***_*m*_ (i.e. all diagonal entries equal to 1). For our studies, we started with annotation scores *A*_*jk*_ for each variant *j* and annotation *k*. We first scaled this value to 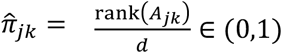, where *d* is the number of variants. This can be interpreted as the probability that a variant is causal based on its annotation score. Variants with larger annotation scores will have *rank*(*A*_*jk*_) closer to *M*, 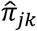 closer to 1. Then the penalty weights were computed as 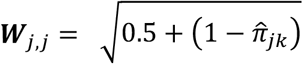, so that variants more likely to be causal are given a smaller penalty. The 0.5 additive constant serves two practical purposes: (1) weights too close to 0 can lead to instabilities within the optimization related to matrix inversion, and (2) the variant with a “median” annotation would have an effective “median” penalty weight of 1. Specifically, we use a group-wise coordinate descent optimization to solve this regression^29,78^:

<u>Step 0</u>: Compute largest eigenvalue for each mask: 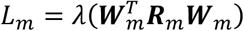, where ***R*** is the submatrix of ***R*** for mask *m*.

<u>Step 1</u>: Marginal correlations for mask *m*:

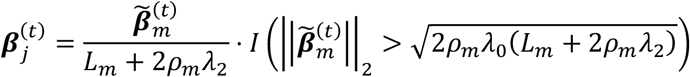

<u>Step 2</u>: Apply group penalty to mask *m*:

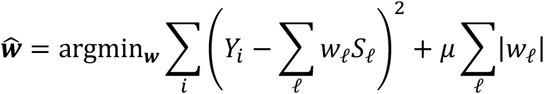

### Ensemble learning

Using individual-level data from an independent tuning dataset, we compute tuning PRS for each method: 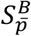 for burden with different p-value thresholds 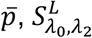 for single-variant L0L2 with parameters *λ*_0_ and *λ*_2_, and 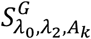 for group L0L2 with parameters *λ*_0_ and *λ*_2_ and functional annotation *A*_*k*_. Especially with a large number of functional annotations, the total number of candidate models can be very large, so we first conduct correlation screening to remove highly correlated candidate models. In this work, we computed the pairwise correlation between all tuning PRS and sequentially removed candidate models with correlation above 0.95. Then, we used 3-fold cross-validation Lasso using glmnet^79^ to combine the remaining candidate models. For the set of all remaining candidate models *S*_*𝓁*_ with *𝓁* = 1, …, *L*, we obtain ensembling weights 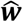 using the penalized regression:

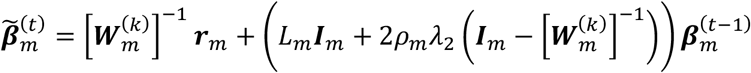

The ensembling weights can then be used to combine the effect estimates from each candidate model 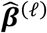 to give the final rare variant effect estimates 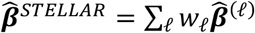.

### Benchmarking STELLAR and incorporating common variants

For our simulation and real data analyses, our primary benchmark was cvPRS, the common variant-only PRS from ALL-Sum. For common variant PRS tuning, we conducted the ensemble learning with respect to outcome residuals, after regressing out covariates (e.g. age, sex, and PCs).

Our other comparator was a “cvPRS+Burden” model that combines cvPRS with burden-based rvPRS. Specifically, we applied 3-fold cross-validation Lasso on the five candidate burden PRS models, adjusted for cvPRS. With either specification of association statistics, the ensembling was always done using outcome residuals after regressing out baseline covariates and the ALL-Sum PRS. Similarly, for STELLAR, we applied the single-variant and group L0L2 penalties to single-variant association statistics, and conducted ensembling on outcome residuals after regressing out baseline covariates and the ALL-Sum PRS.

For final evaluations, we applied each PRS to an independent validation dataset. We evaluated the R^2^ or AUC of the combined ALL-Sum and rare variant PRS within the validation dataset as the main prediction accuracy metrics. For risk stratification, we divided the common and rare variant PRS distributions into deciles for continuous traits and quintiles for disease traits. We then ran regressions of the outcome on the percentile group indicator relative to a reference group (40-60%) that represents “median risk”. The regression coefficients on the group indicator then corresponds to the average change in outcome or odds ratio of disease for each percentile relative to the reference group.

### Evaluation of PRS prediction accuracy

We used partial R^2^ and partial AUC as our key metrics to evaluate PRS accuracy while adjusting for baseline covariates, thus providing a distilled measure of PRS prediction.

For continuous traits, we used the sensemakr package for partial R^2^. In this framework, we first ran a full linear regression with PRSs and covariates:

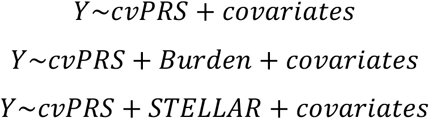

We then used the partial_r2() function to compute the univariate partial R^2^ for cvPRS and group partial R^2^ of cvPRS and rvPRS.

For binary traits, we used the ROCnReg package. Specifically, we used the AROC.sp(), which is a semi-parametric method for covariate-adjusted AUROC. This function can only take one key variable as input, unlike sensemakr which can jointly evaluate cvPRS and rvPRS. Thus, when evaluating the combined prediction of cvPRS and rvPRS, we first ran a multivariate logistic regression (below)

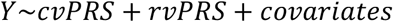

and computed the combined PRS as

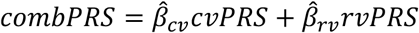

### Importance metrics for selected masks

Using gene-mask information within our method not only enhances prediction by providing structure within the effect estimation but also provides interpretability for the final rare variant PRS. In particular, STELLAR provides mask-level importance metrics, which can be used to identify genes and masks that contribute most to overall and individual-level risk. The importance metrics are computed as follows

The primary metric is the total mask contribution (*T*_*m*_), defined as the sum of squared effects in the final ensembled model: 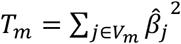. Regardless of the effect direction, this metric describes the overall contribution of the mask to genetic risk, where a high *T*_*m*_ can be driven by a few large effects or several small effects. The second metric is the average mask contribution (*A*_*m*_), intuitively defined as 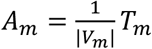. This instead describes a “per-variant” contribution within each mask, which would prioritize masks with consistently large effects, regardless of the mask size. For instance, large masks with only a few large effects may have higher *T*_*m*_ but lower *A*_*m*_. Together, these metrics can offer biological insights into risk-associated genes and may be used to tailor interventions depending on the rare variants an individual carries.

### Whole-exome sequencing data from UK Biobank

We used PLINK format files for WES data of UK Biobank participants (UK Biobank Field #23158). Quality control measures were performed in the following steps. We first removed variants with Hardy-Weinberg Equilibrium p < 1 × 10^−15^. Then, we removed variants for which more than 10% of all genotypes for that variant had a read depth < 10. We finally excluded variants with over 10% missing genotypes and samples with 10% missing genotypes.

### Simulation studies

For our simulation studies, we used observed genotypes for chromosome 19 in the UK Biobank. We used HapMap3 and MEGA reference panels for common variants (MAF ≥ 1%), and WES for rare variants (MAF ≤ 1%). focusing on individuals of European ancestry to avoid population-based confounding. We used 97,820 samples for “training” to compute association summary statistics for common and rare variants, 15,000 samples for “tuning” to conduct ensembling and compute LD for common and rare variants, and 15,000 samples for “validation” to evaluate and compare predictions from different PRS models.

We simulated continuous phenotypes from the observed genotypes using simulated effect sizes with varying proportions of causal variants, effect sizes, and effect directions. In particular, common variant effects were simulated from a normal distribution 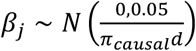, with л_*causal*_ set to 10%, that corresponds to a heritability of 5%. For the rare variants, we first generated synthetic annotations from a 5-dimensional standard multivariate normal distribution, and randomly selected 3 or 5 genes as causal. Then for each gene, we define *K*_0_ as a random subset of 3 out of the five annotations, which were used to determine whether a variant was causal or not:

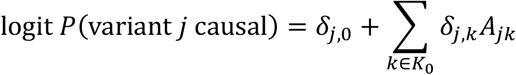

We chose *δ*_*j,k*_ = log (5) and the following values for *δ*_*j*,0_ with corresponding causal probabilities for each functional category (only allowing LOF, DMIS, MIS, and SYN masks to be causal), based on previous simulation designs from STAAR^11^ and SAIGE-GENE+^13^:

| Setting | Quantity | LOF | DMIS / MIS | SYN |
| --- | --- | --- | --- | --- |
| 1 | $\delta_{j,0}$ | logit(0.18) | logit(0.018) | logit(0.0018) |
|  | Causal Prob. | 30% | 10% | 3% |
| 2 | $\delta_{j,0}$ (Rare) | logit(0.18) | logit(0.018) | logit(0.0018) |
|  | Causal Prob. | 30% | 10% | 3% |
| | $\delta_{j,0}$ (Ultrarare) | logit(0.54) | logit(0.054) | logit(0.0054) |
|  | Causal Prob. | 50% | 20% | 5% |
| 3 | $\delta_{j,0}$ | logit(0.06) | logit(0.018) | logit(0.0018) |
|  | Causal Prob. | 20% | 6% | 1% |
| 4 | $\delta_{j,0}$ (Rare) | logit(0.06) | logit(0.018) | logit(0.0018) |
|  | Causal Prob. | 20% | 6% | 1% |
| | $\delta_{j,0}$ (Ultrarare) | logit(0.18) | logit(0.018) | logit(0.0018) |
|  | Causal Prob. | 30% | 10% | 3% |

In settings 2 and 4, we applied separate quantities for rare and ultrarare variants, under an assumption that ultrarare variants may be more likely to contribute to disease than non-ultrarare^80^.

Among causal variants, we then assigned effect sizes according to MAF: weak effects 0.15| log_20_ *MAF* | or strong effects 0.25| log_20_ *MAF* |. These settings correspond to an assumption that rarer causal variants are expected to have stronger effect^80^. Here we considered “Baseline” settings for which all causal variant effects followed the same MAF-based formula, as well as “LOF Boosted” settings, where causal LOF variants have additionally stronger effects than causal DMIS, MIS, and SYN variants. We also further varied the effect directions within causal masks, such that effects were either 100-0, 80-20, or 50-50 in the same direction^18^.

### Real data analysis in UK Biobank

We analyzed 8 continuous traits – LDL, HDL, logTG, TC, height, BMI, EGFR using the CKD-EPI 2021 creatinine-based equation^81^, and HbA1c – and 5 disease outcomes – T2D, CAD, asthma, breast cancer, and prostate cancer. For individuals with reported statin usage, we divided LDL cholesterol measurements by 0.7 and total cholesterol by 0.8, per standard practices^82^. EGFR was computed using creatinine and cystatin measurements according to the CKD-EPI 2021 equation^81^. Our analysis focused on 310,831 individuals of European ancestry in the UK Biobank, with 270,831 individuals used for “training” to compute summary statistics, 20,000 for “tuning”, and 20,000 for “validation”. These individuals were selected according to principal component (PC)-predicted ancestry based a random forest classification model trained in the 1000 Genomes Project Phase 3^83^, where we filtered for individuals with probability of European ancestry greater than 90%. We used 1,546,685 common variants based on HapMap3^30^ and MEGA^31^ arrays, as well as 8,580,612 rare variants from UKBB WES data. We collapsed all ultrarare variants (MAC < 10) into a burden within each mask and treated that as a single variant, giving 1,567,289 effective rare variants.

For the common variants component, we used summary statistics from published consortium-based GWAS: Global Lipids Genetics Consortium for LDL, HDL, logTG, and TC^84^, the Genetic Investigation of ANthropometric Traits (GIANT) consortium for Height and BMI^85,86^, the CKDGen consortium for EGFR^87^, Meta-Analyses of Glucose and Insulin-related traits Consortium (MAGIC) for HbA1C^88^. For binary traits, we obtained GWAS summary statistics from the Transnational-National Asthma Genetic Consortium (TNAGC) ^89^, Coronary Artery Disease Genome-wide Replication and Meta-analysis plus The Coronary Artery Disease Genetics (CARDIoGRAMplusC4D)^90^, Diabetes Genetics Replication And Meta-analysis consortium (DIAGRAM)^59^, Breast Cancer Association Consortium (BCAC) ^91^, and Prostate Cancer Association Group to Investigate Cancer Associated Alterations in the Genome (PRACTICAL)^92^.

We used real functional annotations computed for the UK Biobank WES data provided within the aGDS data format^11,77^. For the group L0L2 penalized regression, we considered an unweighted model (identity matrix for each weighting matrix *W*_*m*_), weights based on CADD^93^, LINSIGHT^94^, and FATHMM-XF^95^, which primarily characterize variant deleteriousness, as well as 8 annotation PCs that characterize a variety of biological functions^11,20^ (**Supplementary Table 3**).

### External validation in All of Us

We further validated the continuous trait PRSs from our main analyses, which were derived in UK Biobank, in the All of Us v7.1 cohort. For the cvPRS component, we used the “ACAF” variant set, which covers variants with minor allele count > 100 and minor allele frequency > 0.01 in at least one of the six computed ancestry groups. For the rvPRS component, we used the “exome” variant set.

For multi-ancestry validation, we included unrelated individuals and stratified by pre-computed ancestry labels, which were assigned based on genetic similarity to one of six populations (AFR, AMR, EAS, EUR, MID, and SAS) in a combined reference dataset of Human Genome Diversity Project and 1000 Genomes Project.

### Quantifying risk stratification

In addition to overall prediction accuracy measured by R^2^, we also evaluated how well common and rare variants can stratify individual-level risk. This has more direct clinical implications, where we can use individuals’ genetic profiles to assess their risk relative to the general population and prioritize individuals at the highest genetic risk for downstream interventions.

For continuous traits, we divided the cvPRS (ALL-Sum) and cvPRS (STELLAR) distributions into deciles. We set the reference group to be individuals whose common and rare variant PRSs are both in the 40-60% group. For each “test group” of individuals within a combination of common and rare variant PRS deciles, we ran the following linear regression:

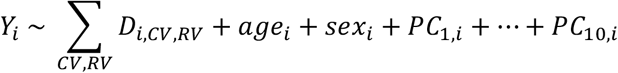

where *D*_*i,CV,RV*_ is a dummy variable that equals 1 if an individual *i* is in common variant percentile *CV* and rare variant percentile *RV*, and 0 for individuals in the reference group. The regression coefficients *β*_*CV,RV*_ for each *D*_*i,CV,RV*_ thus corresponds to the average difference (which we refer to as “relative risk” in the **Results**) in phenotype between the reference and test groups, adjusted for baseline covariates.

For binary traits, due to the relatively small number of cases, we instead divided into quintiles of each PRS distribution. Again, defining the reference group as individuals whose common and rare variant PRSs both fall in the 40-60% group, we ran logistic regressions following the same equation as below. We omitted sex as a covariate for breast cancer and prostate cancer. Exponentiating the regression coefficients *β*_*CV,RV*_ then gives to the odds ratio of disease for individuals in the test group, relative to the reference group, adjusted for covariates. We also repeated marginal regression analyses, one with only cvPRS percentiles 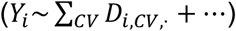 and another with only rvPRS percentiles 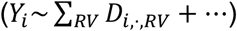, to obtain overall risk stratification for each PRS component on its own.

As a global test for stratification, we employed an inverse-variance weighted test for differences in relative risk estimates *β* (“dBeta”) across all cvPRS percentiles, accounting for standard errors *s* from the above regression analysis). We compared cvPRS only (*β*_*CV*,·_) and cvPRS separated by the *RV* percentile of the STELLAR rvPRS (*β*_*CV,RV*_).

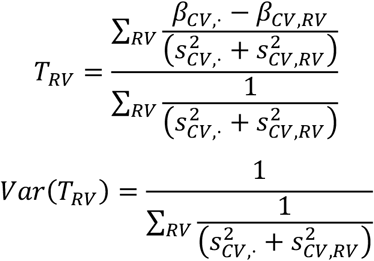

We conducted this test for the top and bottom 10% of the STELLAR rvPRS, using one-sided t-tests corresponding to null hypotheses *H*_0_: *dBeta* = *β*_*CV,RV*_ − *β*_*CV*_ ·≥ 0 for the bottom 10% of rvPRS (i.e., alternative that low rvPRS further reduces risk) and *H*_0_: *dBeta* = *β*_*CV,RV*_ − *β*_*CV*,_ ≤ 0 for the top 10% rvPRS (i.e., alternative that high rvPRS further increases risk).

## Data and Code Availability

Software and tutorials are available at https://github.com/chen-tony/STELLAR. UK Biobank genotype and phenotype data were obtained through application no. 52008. Data from 1000G were obtained from https://www.cog-genomics.org/plink/2.0/resources (genotypes) and https://www.internationalgenome.org/data-portal/sample (population labels). Published GWAS summary statistics for common variants are listed in **Supplementary Table 3D**.

## Acknowledgements

We would like to thank Rui Duan and Peter Kraft for their feedback and insights on this work. We are also grateful to the participants in the 1000 Genomes Project, UK Biobank (resource application no. 52008), All of Us Research Program, and consortium GWAS cohorts for providing vital data to this study. Analysis for this work utilized high-performance computing resources from the Faculty of Arts and Sciences Research Computing Cluster at Harvard University. This research was supported by NIH Training Grant T32GM135117 and NSF Graduate Research Fellowship DGE-2140743 (T.C.), NIH grants 1R01HL173044 and 1R01AG085581 (X.Li). NIH Intramural Research Program (H.Z.), Office of Naval Research grants N000142112841 and N000142212665 (R.M.), and NIH grants R35-3 CA197449, U19-CA203654, R01-HL163560, U01-HG009088, U01-HG012064 (X.Lin).

## Notes

### Competing Interest Statement

The authors have declared no competing interest.

### Author Declarations

UK Biobank: Ethical approval was granted by the National Research Ethics Service Committee North West, Haydock (Reference Number: 11/NW/0382). Detailed information can be found here: https://www.ukbiobank.ac.uk/media/cgjh1ohn/favourable-ethical-opinion-and-rtb-approval-16-nw-0274-200778-may-2016.pdf All of Us: The study was approved by the Institutional Review Board of the All of Us Research Program. Detailed information can be found here: https://allofus.nih.gov/about/who-we-are/institutional-review-board-irb-of-all-of-us-research-program.

