## Supplementary Figures for "STELLAR: A flexible ensemble learning framework integrating rare variants to enhance polygenic risk prediction"

**Supplementary Figure 1. Full comparisons of prediction accuracy in simulation studies.** Using observed genotypes from UK Biobank, we simulated continuous phenotypes with 10% of common variants contributing 5% heritability and varied effect sizes and directions for rare variants. We varied the number of causal genes to be 3 (**A, C**) or 5 (**B, D**), effect sizes to be follow either $\boldsymbol{\beta} = \mathbf{0.25}|\mathbf{log_{10}}\,\boldsymbol{MAF}|$ or $\mathbf{0.15}|\mathbf{log_{10}}\,\boldsymbol{MAF}|$, rare variant causal proportions to be {30% of LOF, 10% of MIS, 3% of SYN} or {20% of LOF, 6% of MIS, 1% of SYN}, and additional scenarios where ultrarare variants (MAC < 10) were twice as likely to be causal (**C**, **D**). Summary statistics were computed from 97,820 UK Biobank samples, PRS were tuned using 10,000 samples and validated in a separate 10,000. We compared a baseline PRS with only common variants (cvPRS) using ALL-Sum, a combined PRS of ALL-Sum and Burden (cvPRS+Burden) scores, and our proposed combination of ALL-Sum and STELLAR (cvPRS+STELLAR). We repeated each simulation setting 50 times, and reported the average $R^2$ for each PRS (text boxes) as well as percentage differences between PRSs with Bonferroni-adjusted p-values for these differences using paired t-tests (floating text), where (**) indicates $p < 0.01$, (*) $p < 0.05$, and (ns) $p > 0.05$.

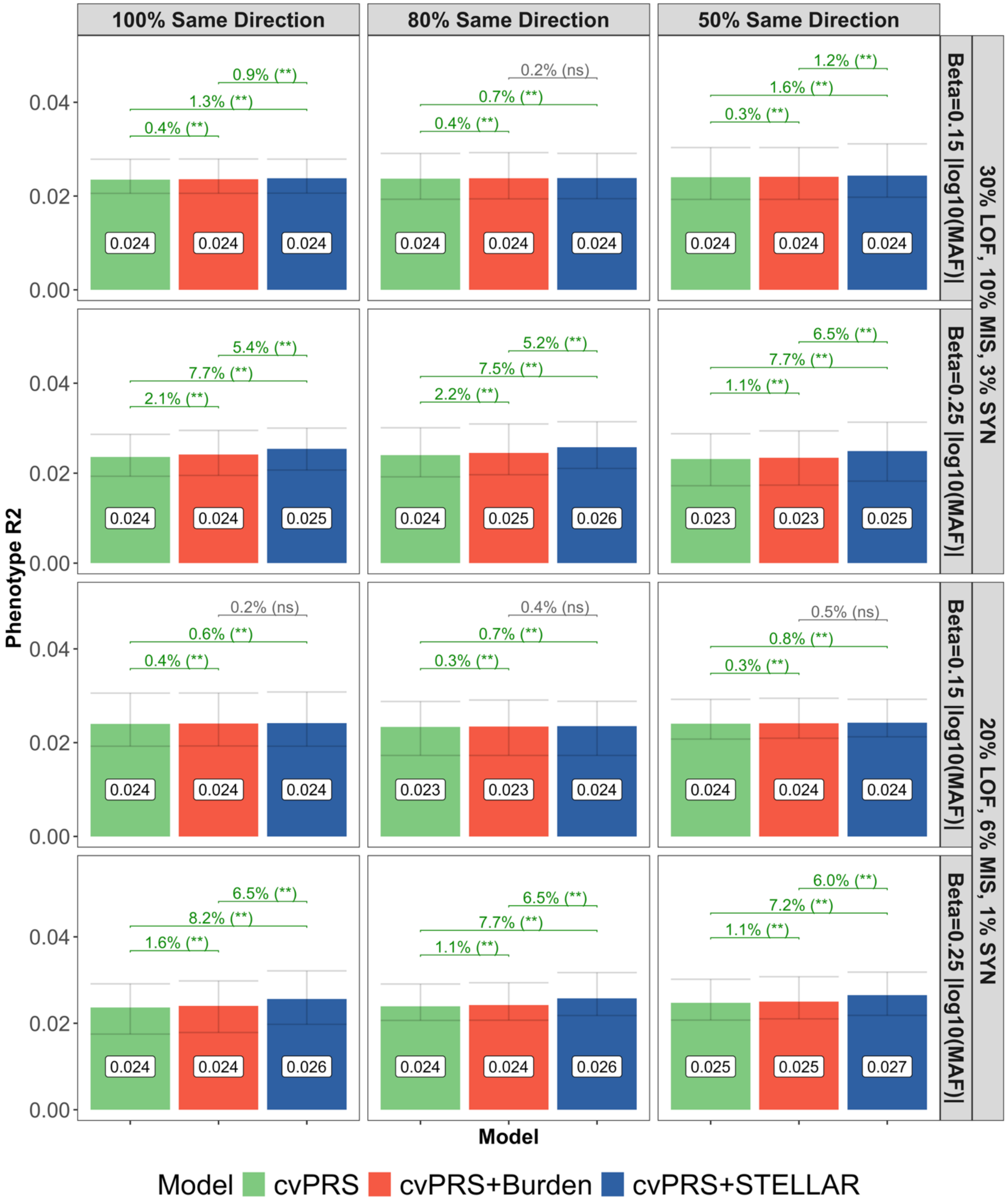


**Supplementary Figure 1A. Simulation results for 3 causal genes.**

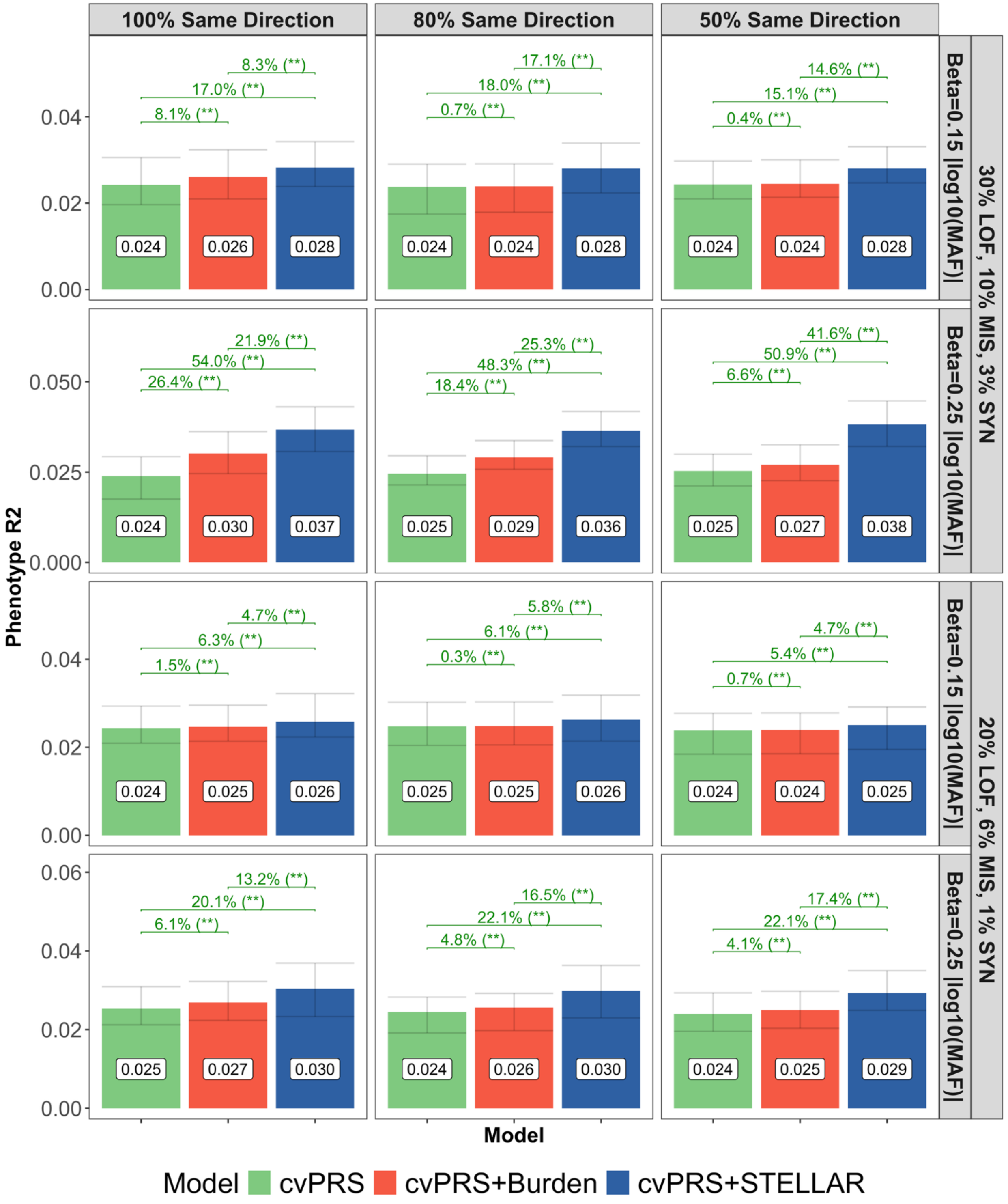


**Supplementary Figure 1B. Simulation results for 5 causal genes.**

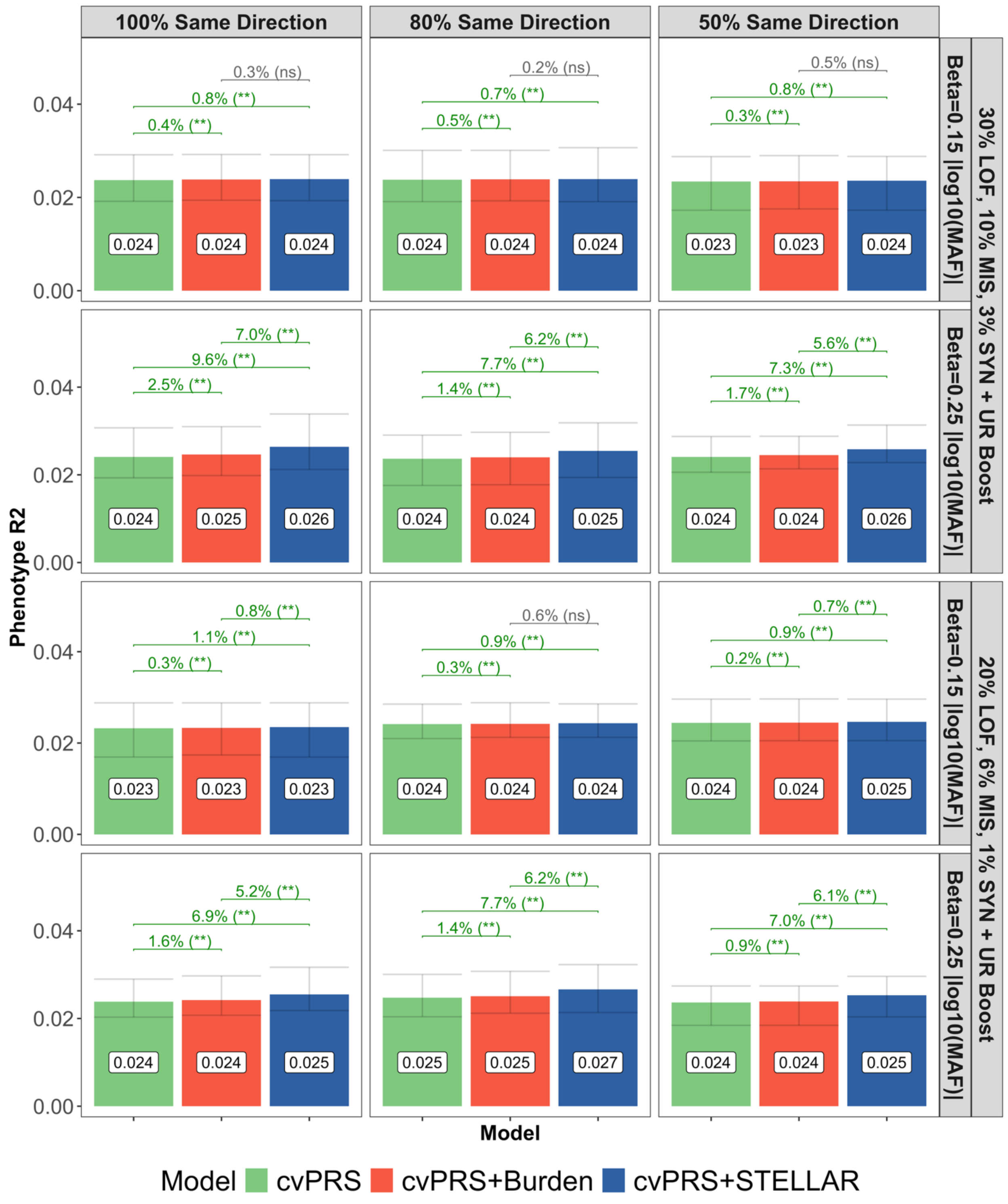


**Supplementary Figure 1C. Simulation results for 3 causal genes, with ultrarare variants more likely to be causal.**

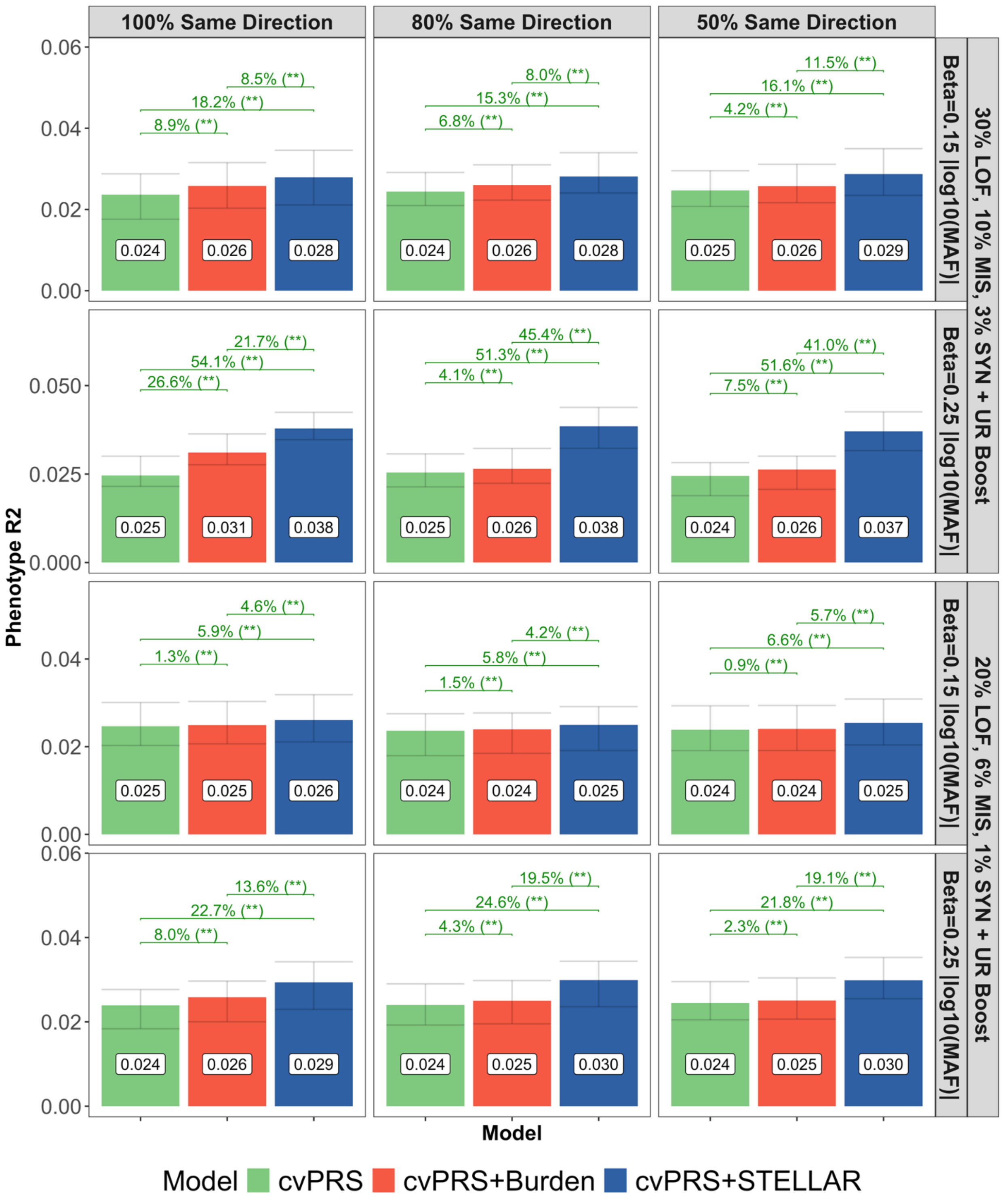


**Supplementary Figure 1D. Simulation results for 5 causal genes, with ultrarare variants more likely to be causal.**

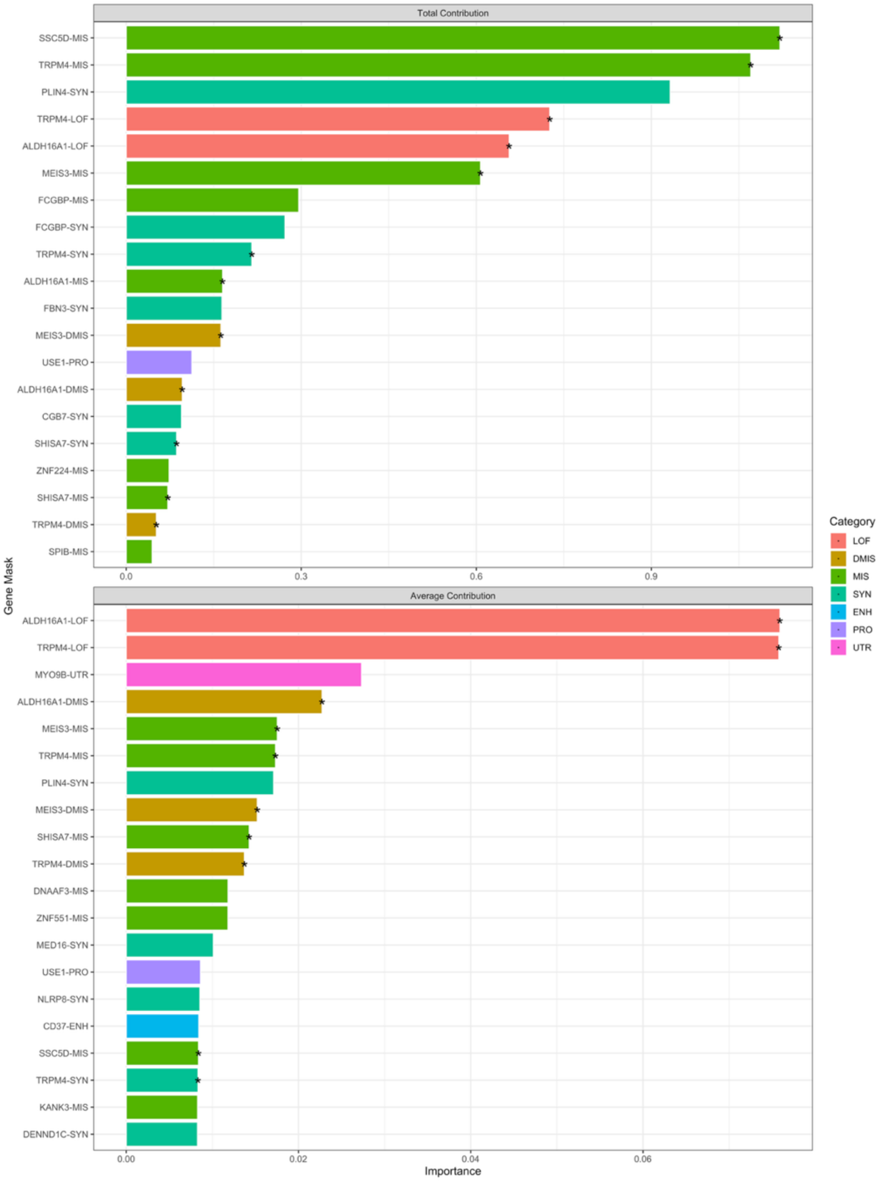


**Supplementary Figure 2. Importance metrics from simulations using 5 causal genes.** We used effect estimates from STELLAR to compute Total Contribution ($\boldsymbol{T_m} = \sum_{j \in V_m} \boldsymbol{\beta_j^2}$) and Average Contribution ($\boldsymbol{A_m} = \frac{\mathbf{1}}{|\boldsymbol{V_m}|} \sum_{j \in V_m} \boldsymbol{\beta_j^2}$) importance metrics. The results shown here correspond to 5 causal genes (30% LoF, 10% missense, and 3% synonymous causal variants, with $\boldsymbol{\beta} = \mathbf{0.25}\,|\,\mathbf{log_{10}}\,\boldsymbol{MAF})|$), 10% causal common variants (5% heritability). Barplots show the top 20 masks for each metric, averaged across the 50 simulation replicates, with stars indicating true causal masks.

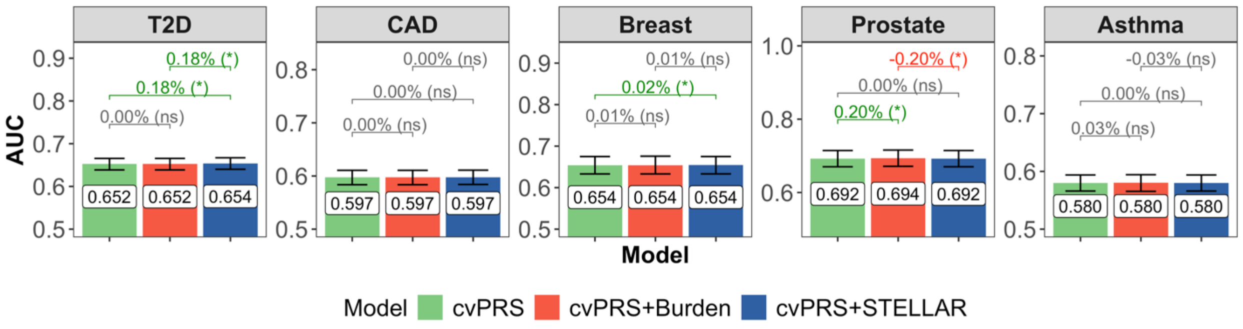


**Supplementary Figure 3. Comparison of AUC between PRS models for 5 binary traits.** We show the observed partial AUC of each PRS with error bars representing the 95% BCI for AUC. We also evaluated the BCI in the percentage difference in AUC between methods, where (*) indicates that the 95% BCI does not include 0 and (ns) indicates that the 95% BCI includes 0.

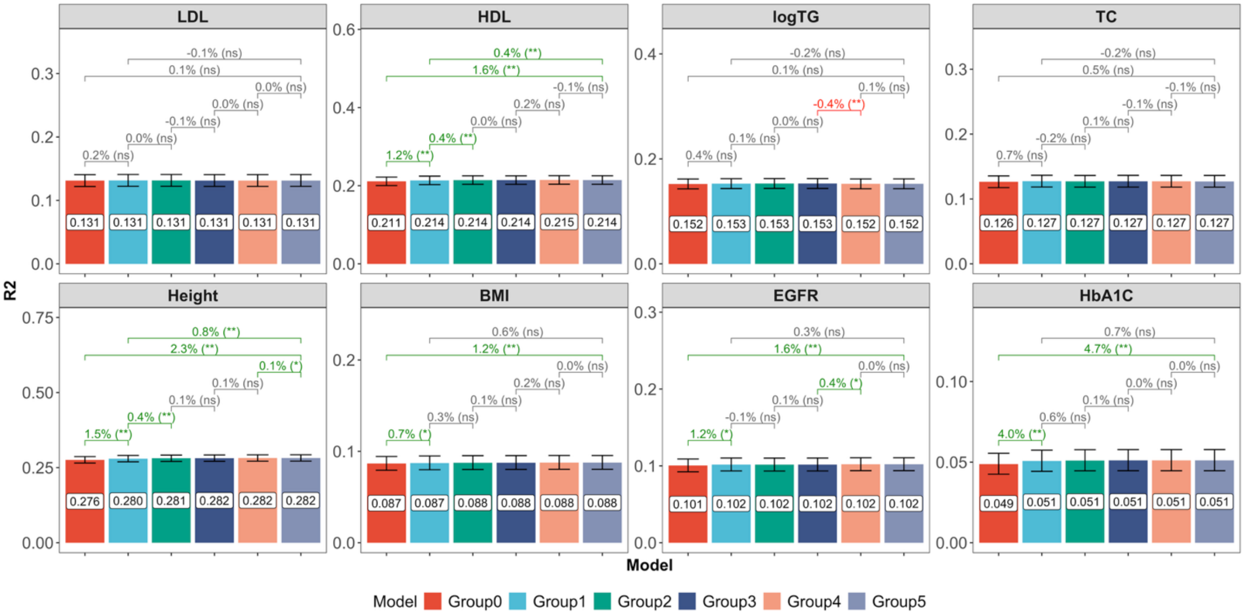


**Supplementary Figure 4. Prediction accuracy of STELLAR with different combinations of functional annotations.** Groups: 0) only Burden and single-variant L0L2; 1) add group L0L2 (no annotation weights); 2) add CADD annotation weights; 3) add LINSIGHT and FATHMM-XF annotation weights; 4) add epigenetic active, epigenetic repressed, epigenetic transcription, and conservation annotation PC weights; 5) add local nucleotide diversity, mappability, transcription factor, and protein function annotation PC weights. We also evaluated the BCI in the percentage difference in $R^2$ between select pairs of methods, where (**) indicates that the 99% BCI does not include 0, (*) that the 95% BCI does not include 0 and (ns) that the 95% BCI includes 0.

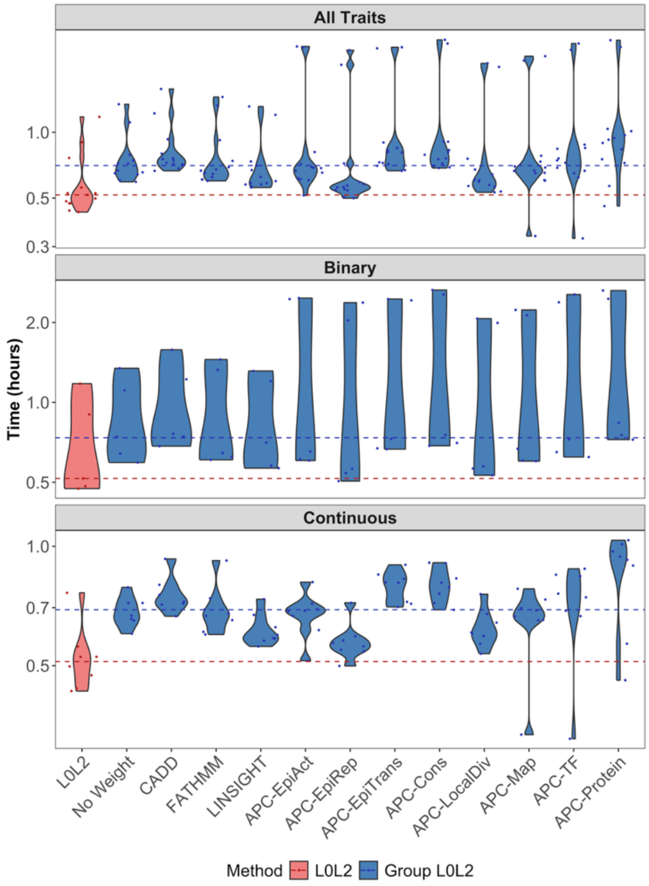


**Supplementary Figure 5. Runtime of penalized regression models with different functional annotations.** We evaluated the runtime of L0L2 and group L0L2 (using twelve different weighting schemes) penalized regression for rare variants across the 8 continuous traits and 5 disease outcomes. We reported the runtime of each model in hours, with horizontal lines indicating the median runtime for L0L2 (red) and all group L0L2 (blue) models.

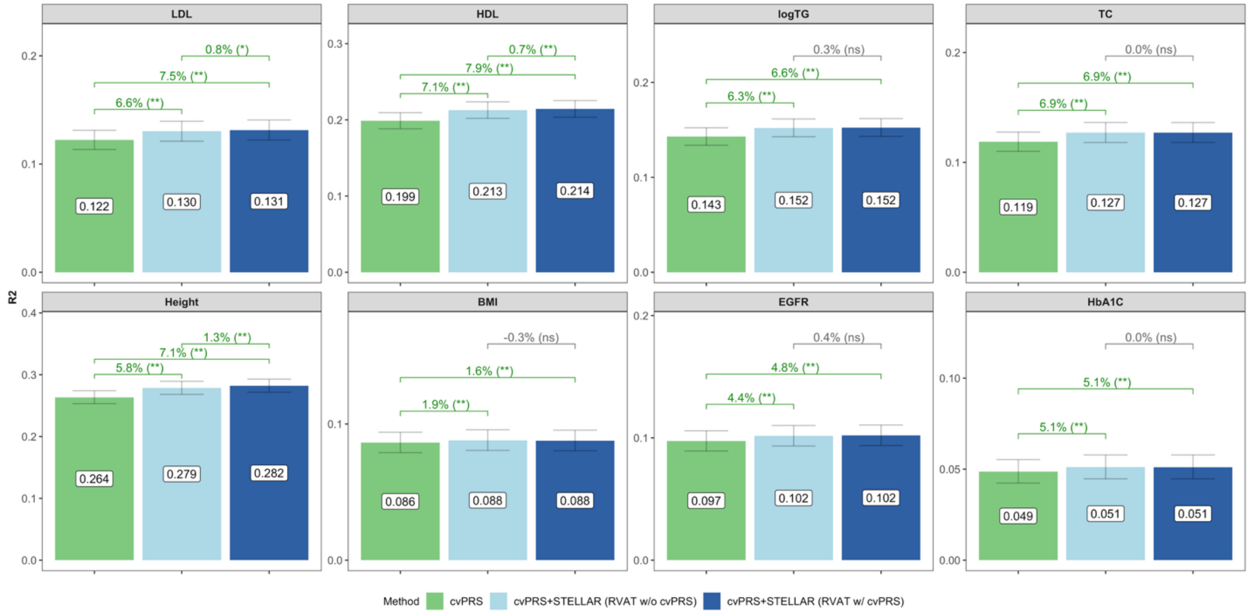


**Supplementary Figure 6. Prediction accuracy with and without adjusting for common variant PRS in rare variant association testing.** We compared the prediction accuracy of STELLAR using rare variant association summary statistics with and without adjusting for the common variant PRS. We show the partial $R^2$ for the common variant PRS (green), STELLAR without adjusting for common variants (light blue), and the main implementation of STELLAR after adjusting for common variants (dark blue). We also evaluated the BCI in the percentage difference in $R^2$ between methods, where (**) indicates that the 99% BCI does not include 0, (*) that the 95% BCI does not include 0 and (ns) that the 95% BCI includes 0.

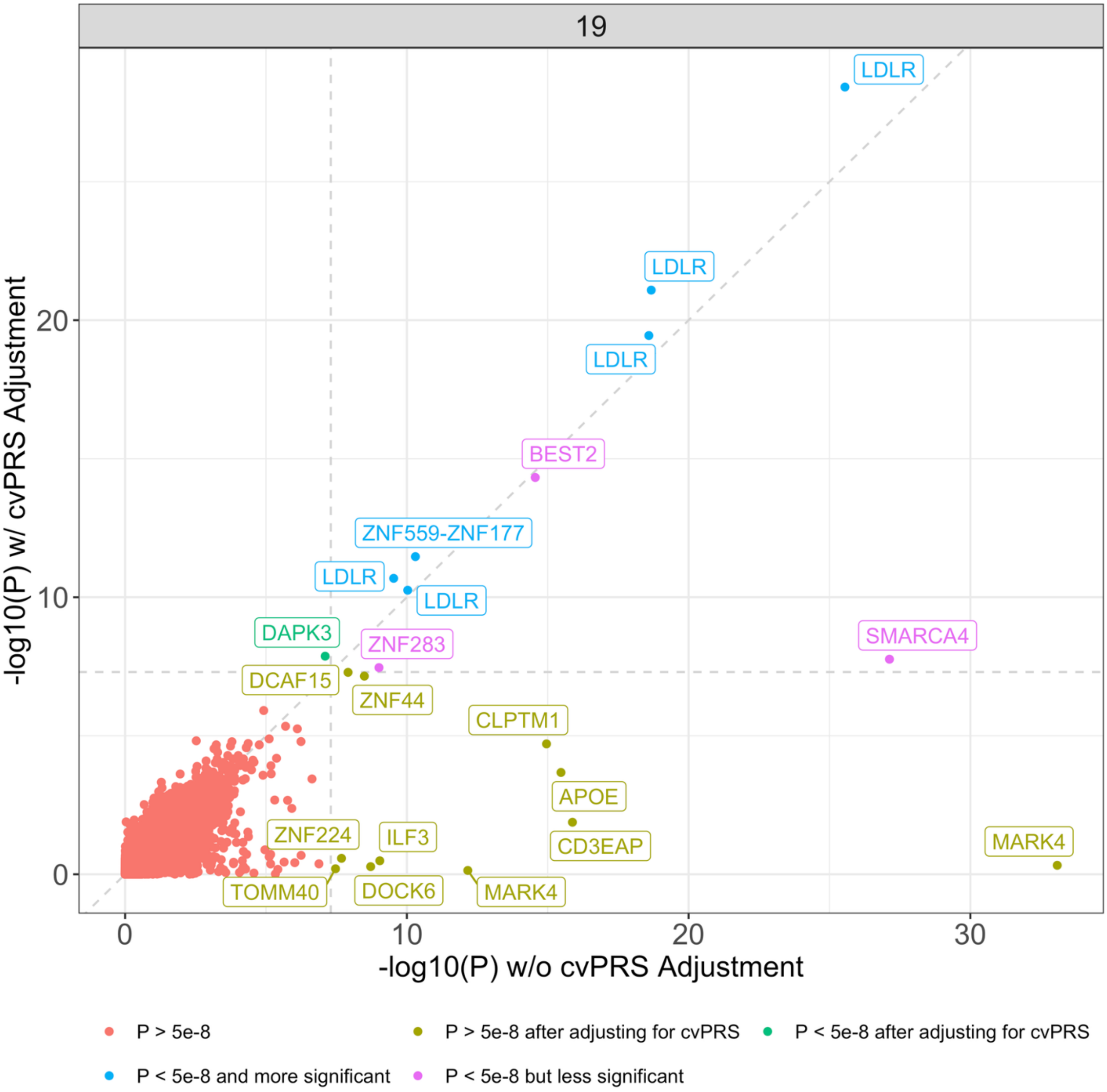


**Supplementary Figure 7. Rare variant association p-values with and without adjusting for common variant PRS.** We compared single-rare-variant and ultrarare-burden association p-values (negative log-10 scale) for LDL on chromosome 19, with and without adjusting for the common variant PRS using ALL-Sum on GLGC-based GWAS summary statistics. Horizontal and vertical dotted lines represent $p=5x10^{-6}$, and the diagonal represents the identity line. We colored points according to different sections of the graph, and annotated points with the genes they are associated with.

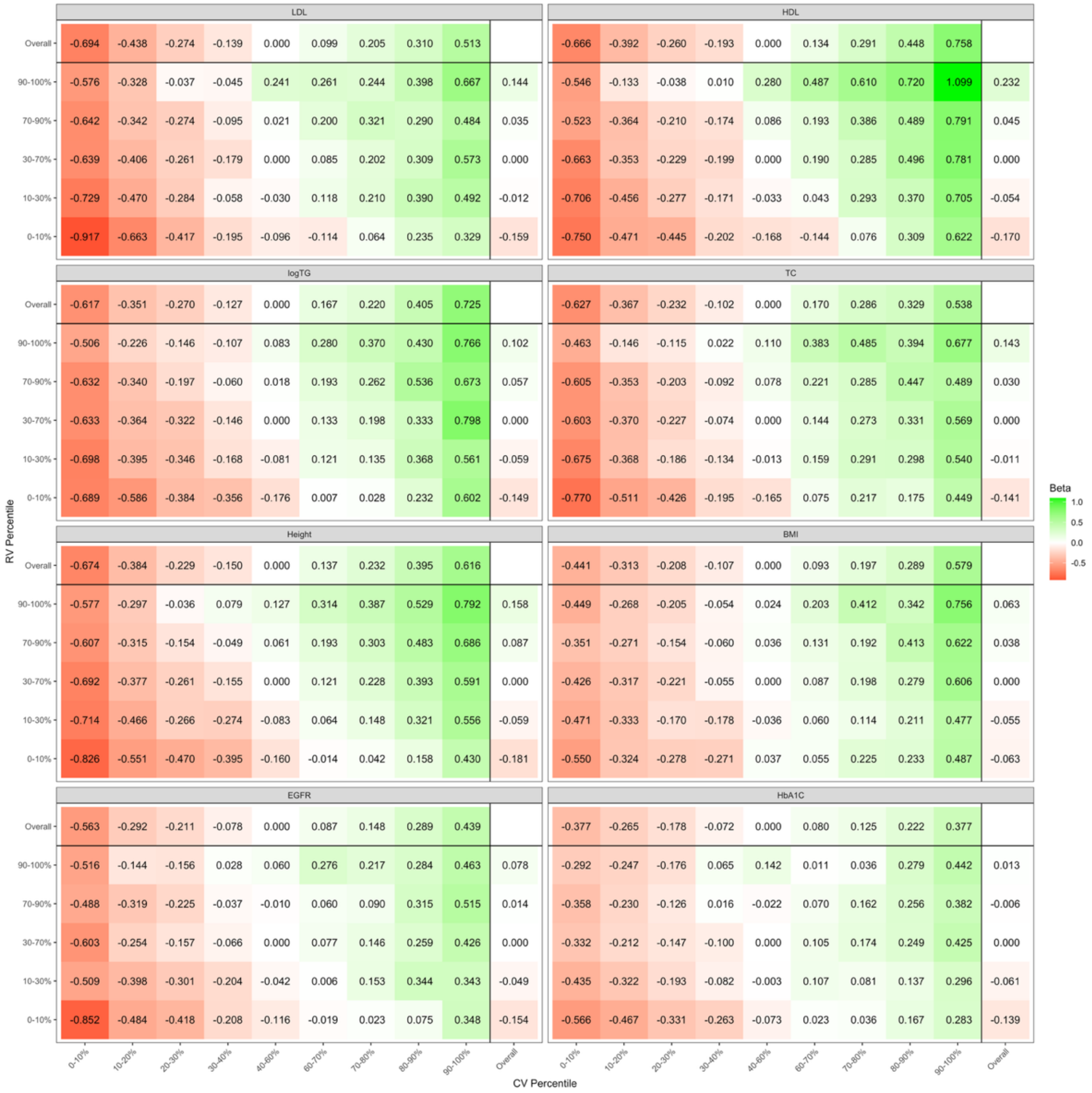


**Supplementary Figure 8. Risk stratification of continuous traits using percentiles of common and rare variant PRS.** We evaluated the change in continuous trait across percentiles of both cvPRS and STELLAR rvPRS. We ran linear regression models of outcome on a group indicator adjusted for age, sex, and PCs, where the resulting coefficient for the group indicator ("Beta" on the y-axis) quantifies the average increase in the outcome in each percentile, relative to the reference group. Here, the reference group was defined as individuals whose rare and common variants were both within the 40-60% percentiles of the respective distributions, for whom "Beta" is just 0. We visualized the "Beta" values across deciles of the cvPRS on the x-axis and STELLAR rvPRS on the y-axis. In each panel, the top row corresponds to "Beta" only looking marginally across cvPRS percentiles, and the rightmost column marginally across rvPRS percentiles.

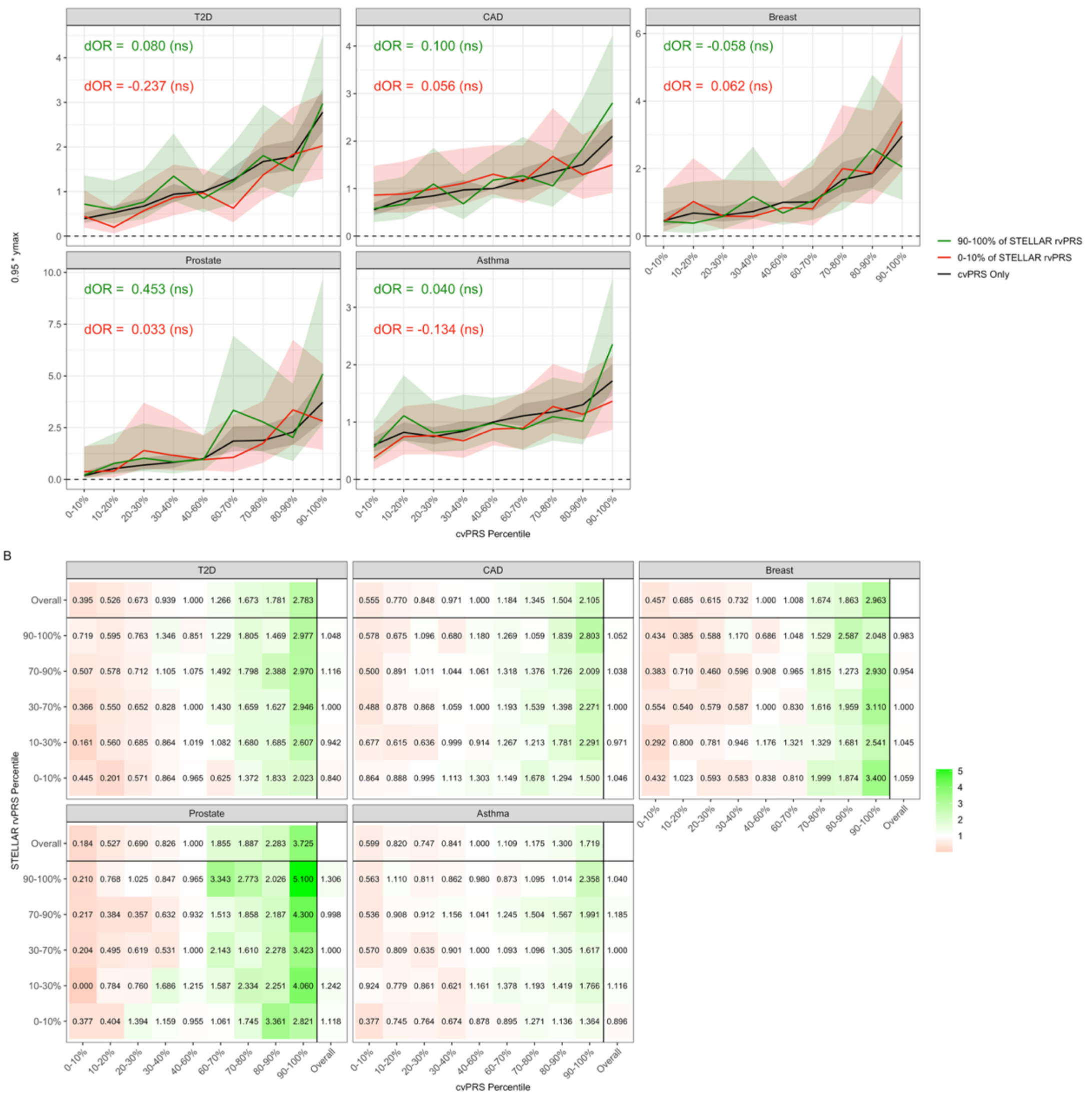


**Supplementary Figure 9. Risk stratification of binary traits using percentiles of common and rare variant PRS.** We evaluated the change in continuous trait across percentiles of both rare and common variants PRSs. We ran logistic regression models of outcome on a group indicator adjusted for age, sex, and PCs, where exponentiating the resulting coefficient for the group indicator quantifies the odds ratio (OR) for disease in each percentile, relative to the reference group. Here, the reference group was defined as individuals whose rare and common variants were both within the 40-60% percentiles of the respective distributions, for whom the OR is 1. (A) Odds ratios with respect to cvPRS percentiles overall and further separated at the top and bottom 10% of the STELLAR rvPRS (lines), 95% confidence intervals for the difference in ORs (ribbons), and average difference in ORs with accompany one-sided p-values (text). (B) Marginal and stratified odds ratios numerically tabulated across all percentiles of cvPRS and STELLAR rvPRS.

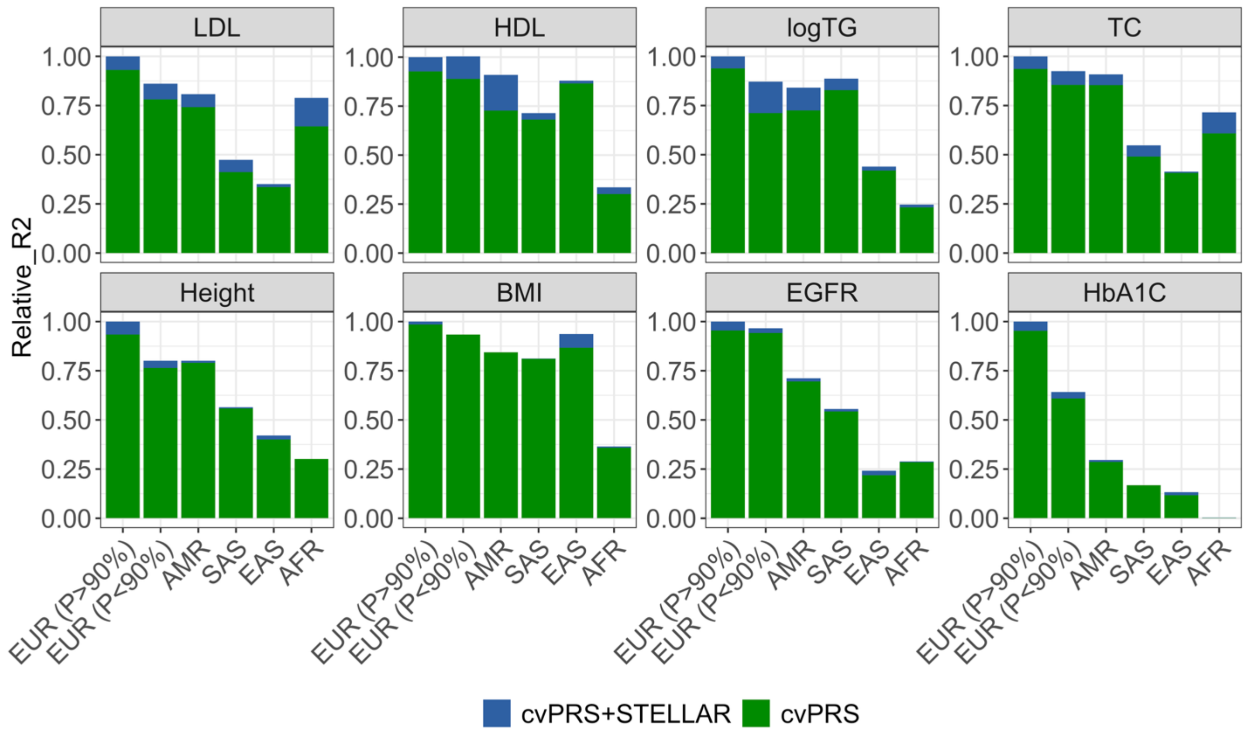


**Supplementary Figure 10. Trait-specific comparison of prediction accuracy across ancestry groups.** We compared PRS prediction accuracy across ancestry groups in the UK Biobank, separated by each of the eight continuous traits. In particular, we evaluated the relative accuracy compared to the cvPRS+STELLAR in European ancestry with predicted ancestry probability greater than 90%, which is most similar to the UKBB training and tuning data. We show the relative $R^2$ for the baseline common variants PRS in green, and the additional $R^2$ from STELLAR in dark blue.

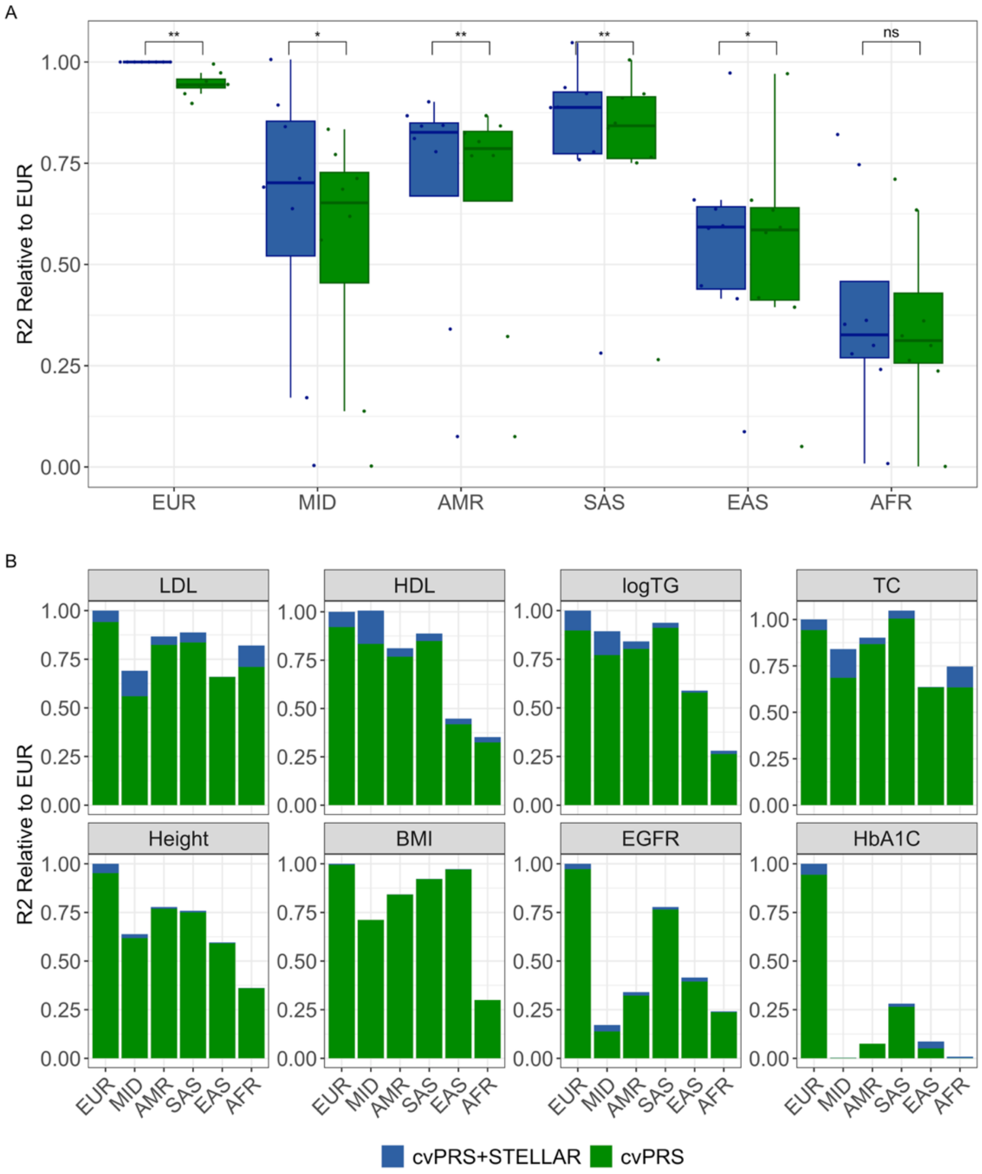


**Supplementary Figure 11. Prediction accuracy across ancestry groups in All of Us.** We evaluated prediction accuracy of the UKBB-based PRSs in six ancestry groups within the AoU cohort. We compared the relative accuracy of each PRS compared to cvPRS+STELLAR in European ancestry. (A) Points and boxplots indicate relative $R^2$ for each of the 8 continuous traits, colored by either cvPRS (green) or cvPRS+STELLAR (blue). We used paired t-tests to compare relative accuracy within each ancestry: (**) $p < 0.01$; (*) $p < 0.05$; (ns) $p > 0.05$. (B) Barplots show the relative accuracy compared to cvPRS+STELLAR in European ancestry. We show the relative $R^2$ for the baseline common variants PRS in green, and the additional $R^2$ from STELLAR in dark blue.

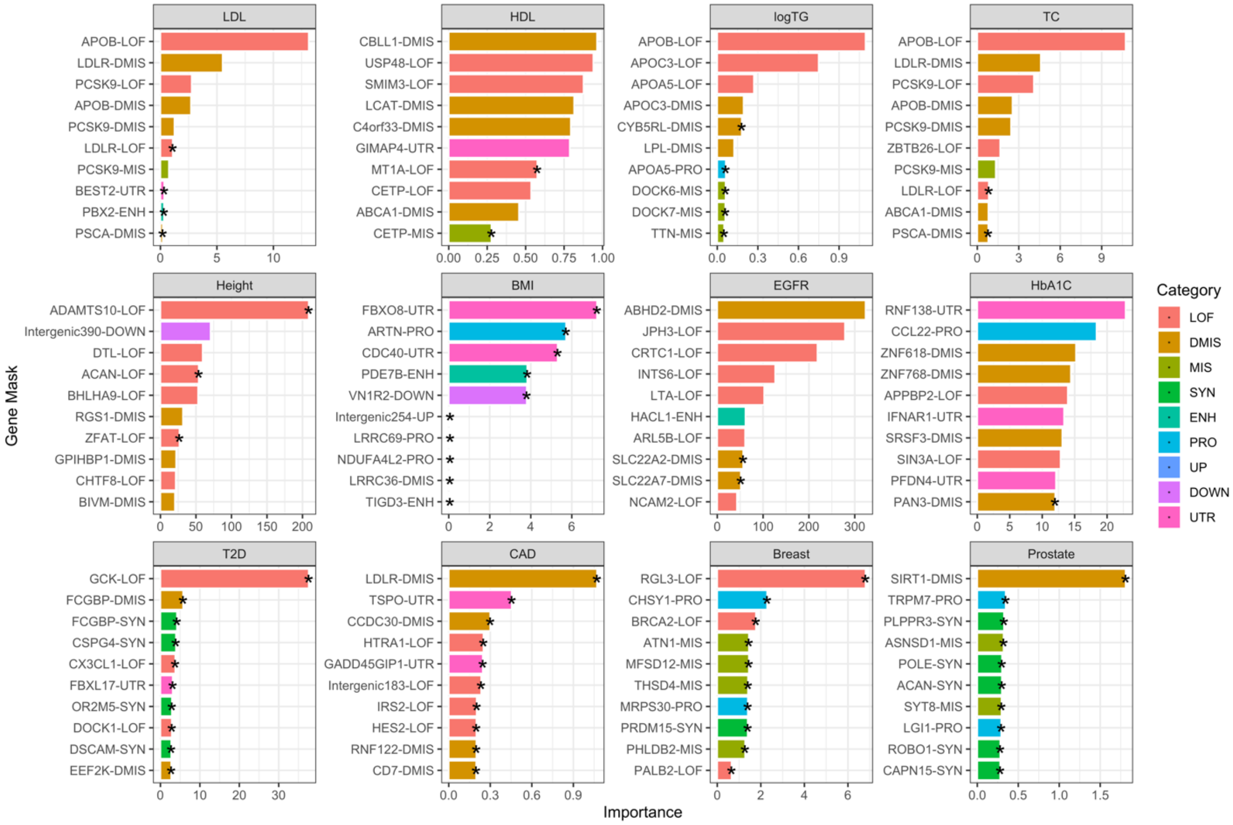


**Supplementary Figure 12. Top ten gene masks for Total Contribution ($T_m$) importance metric using STELLAR.** Using estimated rare variant effect sizes from STELLAR, we computed the total contribution score of each gene mask as $T_m = \sum_{j \in V_m} \beta_j^2$ for each trait. We added stars to denote masks that were not in the top 10 using only Burden models, and thus were uniquely prioritized by STELLAR.

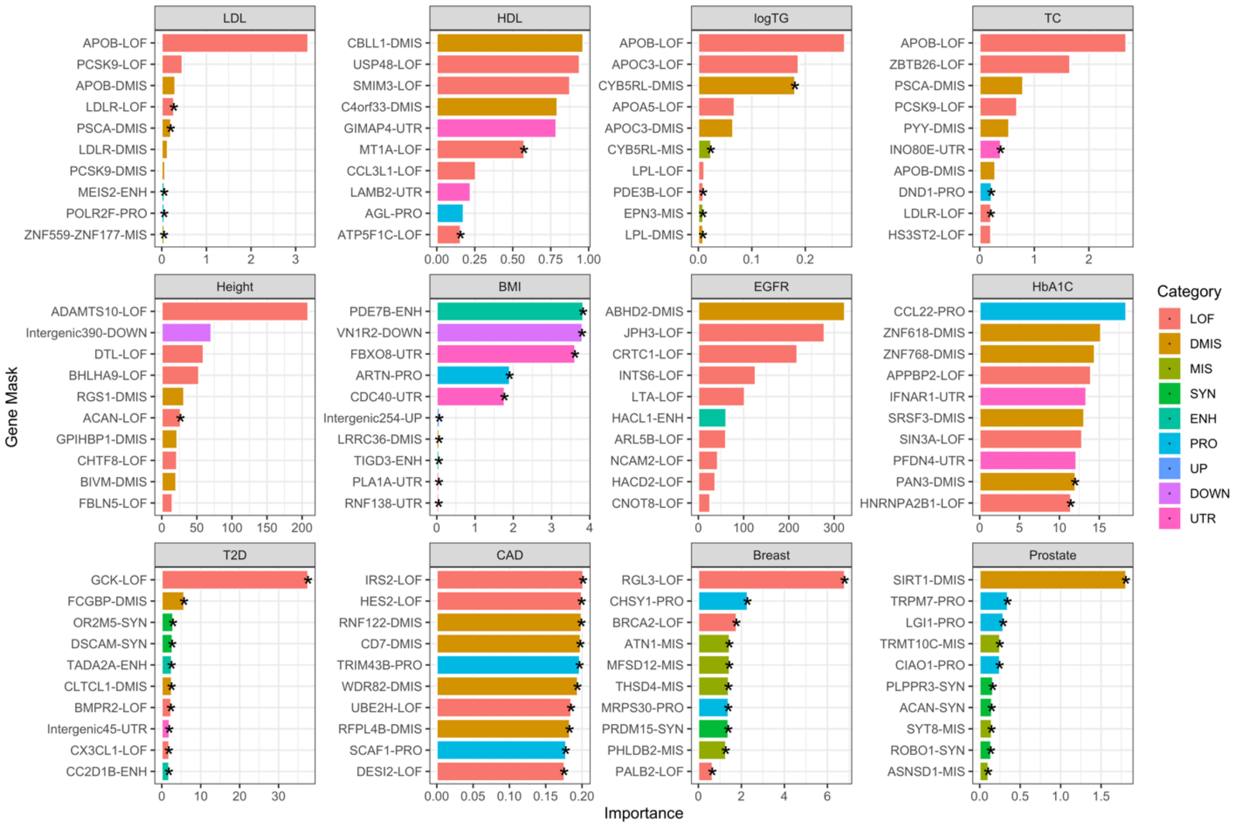


**Supplementary Figure 13. Top ten gene masks for Average Contribution ($A_m$) importance metric using STELLAR.** Using estimated rare variant effect sizes from STELLAR, we computed the average contribution score of each gene mask as $\boldsymbol{A_m} = \frac{\mathbf{1}}{|\boldsymbol{V_m}|}\sum_{\boldsymbol{j} \in \boldsymbol{V_m}} \boldsymbol{\beta_j^2}$ for each trait. We added stars to denote masks that were not in the top 10 using only Burden models, and thus were uniquely prioritized by STELLAR.

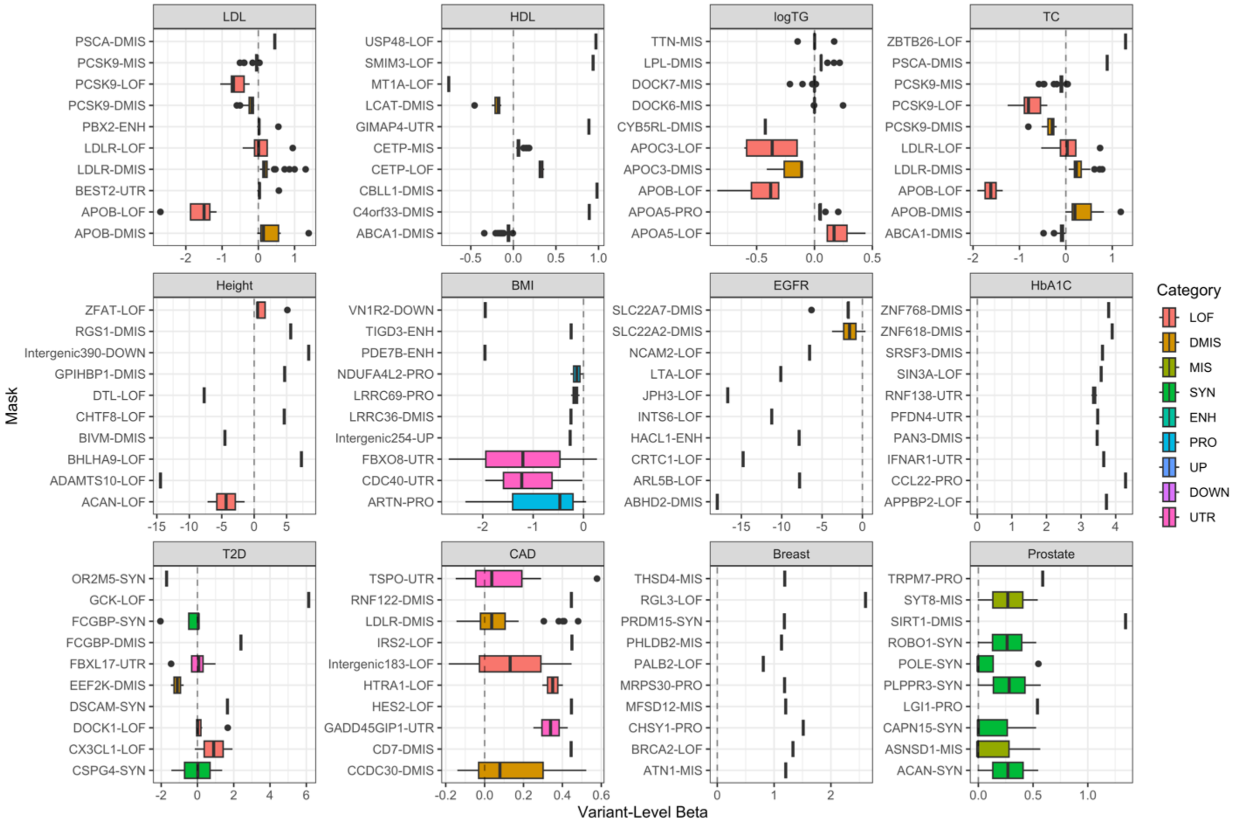


**Supplementary Figure 14. Estimated variant-level effect sizes for top ten masks based on Total Contribution importance.** We visualized the estimated variant- or ultrarare-burden-level effect sizes for gene masks with the top ten total contribution importance metric in each trait. Boxplots are colored by the functional category of the selected mask, with a vertical dotted line at 0 to indicate when effect sizes span both positive and negative values.

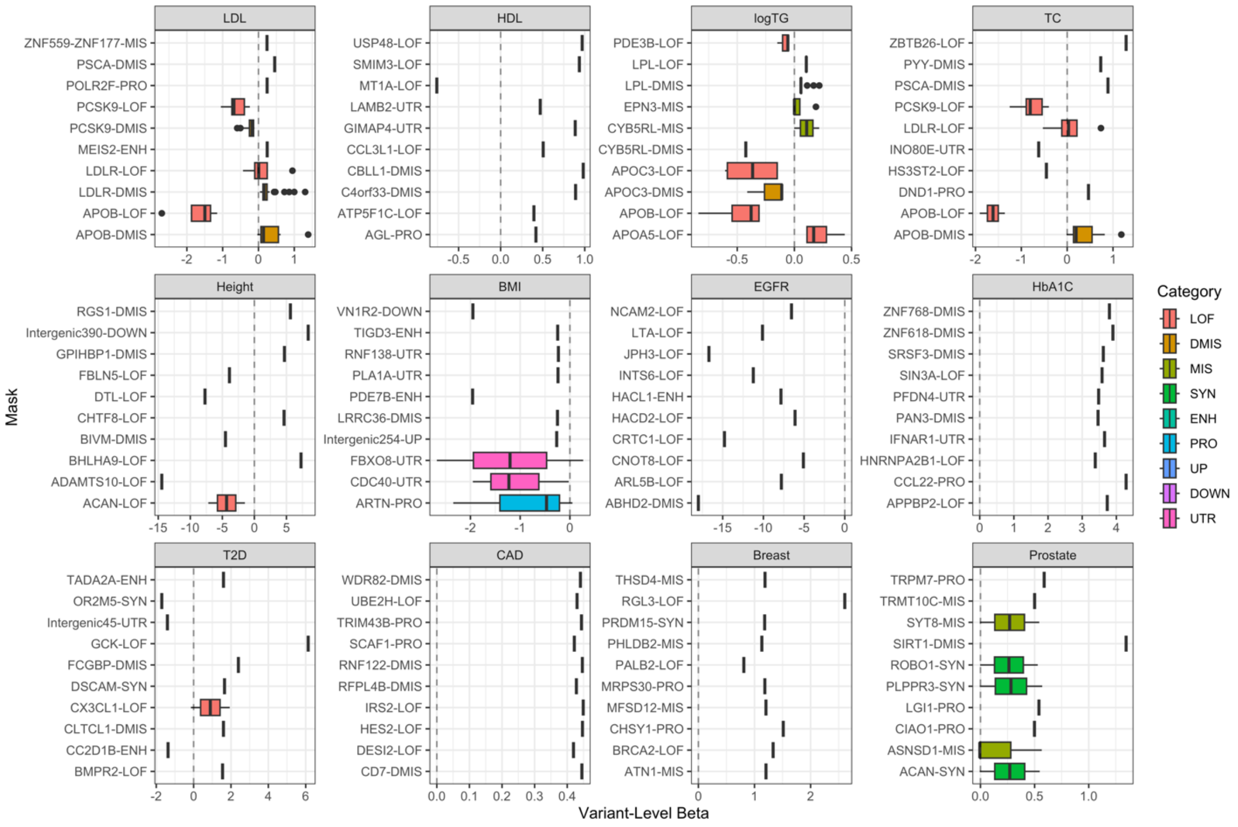


**Supplementary Figure 15. Estimated variant-level effect sizes for top ten masks based on Average Contribution importance.** We visualized the estimated variant- or ultrarare-burden-level effect sizes for gene masks with the top ten average contribution importance metric in each trait. Boxplots are colored by the functional category of the selected mask, with a vertical dotted line at 0 to indicate when effect sizes span both positive and negative values.
